# Impaired immune signaling and changes in the lung microbiome precede secondary bacterial pneumonia in COVID-19

**DOI:** 10.1101/2021.03.23.21253487

**Authors:** Alexandra Tsitsiklis, Beth Shoshana Zha, Ashley Byrne, Catherine DeVoe, Elze Rackaityte, Sophia Levan, Sara Sunshine, Eran Mick, Rajani Ghale, Christina Love, Alexander J. Tarashansky, Angela Pisco, Jack Albright, Alejandra Jauregui, Aartik Sarma, Norma Neff, Paula Hayakawa Serpa, Thomas J. Deiss, Amy Kistler, Sidney Carrillo, K. Mark Ansel, Aleksandra Leligdowicz, Stephanie Christenson, Angela Detweiler, Norman G. Jones, Bing Wu, Spyros Darmanis, Susan V. Lynch, Joseph L. DeRisi, COMET Consortium, Michael A. Matthay, Carolyn M. Hendrickson, Kirsten N. Kangelaris, Matthew F. Krummel, Prescott G. Woodruff, David J. Erle, Oren Rosenberg, Carolyn S. Calfee, Charles R. Langelier

## Abstract

Secondary bacterial infections, including ventilator-associated pneumonia (VAP), lead to worse clinical outcomes and increased mortality following viral respiratory infections including in patients with coronavirus disease 2019 (COVID-19). Using a combination of tracheal aspirate bulk and single-cell RNA sequencing we assessed lower respiratory tract immune responses and microbiome dynamics in 23 COVID-19 patients, 10 of whom developed VAP, and eight critically ill uninfected controls. At a median of three days (range: 2-4 days) before VAP onset we observed a transcriptional signature of bacterial infection. At a median of 15 days prior to VAP onset (range: 8-38 days), we observed a striking impairment in immune signaling in COVID-19 patients who developed VAP. Longitudinal metatranscriptomic analysis revealed disruption of lung microbiome community composition in patients with VAP, providing a connection between dysregulated immune signaling and outgrowth of opportunistic pathogens. These findings suggest that COVID-19 patients who develop VAP have impaired antibacterial immune defense detectable weeks before secondary infection onset.

## Introduction

Secondary bacterial pneumonia results in significant morbidity and mortality in patients with viral lower respiratory tract infections (LRTI)^1^. This problem was evident in the 1918 influenza pandemic during which the majority of deaths were ultimately attributed to secondary bacterial pneumonia^2^. SARS-CoV-2 infection, like influenza, confers an increased risk of late onset secondary bacterial infection, often manifesting as ventilator-associated pneumonia (VAP)^3, 4^. Marked heterogeneity exists, however, with respect to the risk of VAP in patients with coronavirus disease 2019 (COVID-19), with incidence ranging from 12-87% in published cohort studies^5–8^.

The mechanisms underlying VAP susceptibility in COVID-19 remain unknown. Animal models of influenza may provide some insight, suggesting a role for interferon-mediated suppression of cytokines essential for bacterial defense, including those important for neutrophil recruitment, antimicrobial peptide production and the Th17 response^9–11^. The impact of interferon signaling on antibacterial host defense, however, varies based on bacterial pathogen, and is not well studied in humans^12^.

Much of our understanding of VAP immunobiology derives from sepsis studies, which describe a state of immune reprogramming in patients who go on to develop nosocomial infections^13–16^. This state is characterized by impaired monocyte, neutrophil and CD4 lymphocyte function, as well as decreased responsiveness of pathogen recognition receptors and downstream signaling pathways^13–16^. Severe COVID-19, like sepsis, is characterized by a dysregulated host response to infection, raising the possibility that similar mechanisms may contribute to the development of VAP in patients infected with SARS-CoV-2.

In addition to host immune elements, microbial factors contribute to VAP pathogenesis. Mechanically ventilated patients endure prolonged exposure to the hospital environment, which increases risk of colonization by opportunistic pathogens in the upper airway, oropharynx, and lungs^14, 16^. Micro-aspiration or direct invasion may subsequently lead to lower respiratory tract disease and clinical onset of VAP^16, 17^. Further, the common practice of early empiric antimicrobial administration in critically ill patients can lead to disruption of the lung microbiome, which has been associated with the development of VAP^18, 19^.

Despite their interconnected roles, no studies to date have simultaneously profiled host immune responses and lung microbiome dynamics in the context of VAP. Further, the question of whether host responses to SARS-CoV-2 or other viral infections may contribute to this dysbiosis, leading to VAP, remains unanswered.

Given the marked heterogeneity in VAP incidence among patients with COVID-19^5–8^, as well as gaps in mechanistic understanding of secondary bacterial pneumonia, we sought to assess the molecular determinants of VAP in patients infected with SARS-CoV-2. We employed a systems biology approach involving immunoprofiling the host transcriptional response and simultaneously assessing lung microbiome dynamics, using a combination of bulk and single cell RNA sequencing and extensive clinical phenotyping. We observed a striking impairment in antibacterial immune signaling at the time of intubation, that correlated with disruption of the lung microbiome, weeks before the onset of VAP.

## Results

We conducted a prospective case-control study of adults requiring mechanical ventilation for COVID-19 or for illnesses other than pneumonia. Of 63 patients with COVID-19 initially enrolled, tracheal aspirate (TA) specimens from 23 patients met inclusion criteria for primary analysis (**Methods,** **Figure 1**). In addition, eight critically ill patients without COVID-19 from a second cohort (Study 2, **Methods**) were included as controls. Patients were enrolled at one tertiary care hospital and one safety net hospital in San Francisco, California under research protocols approved by the University of California San Francisco Institutional Review Board **(Methods)**. We collected TA periodically following intubation and performed bulk and single cell RNA-seq (scRNA-seq) **(Methods)**.

**Figure 1:**
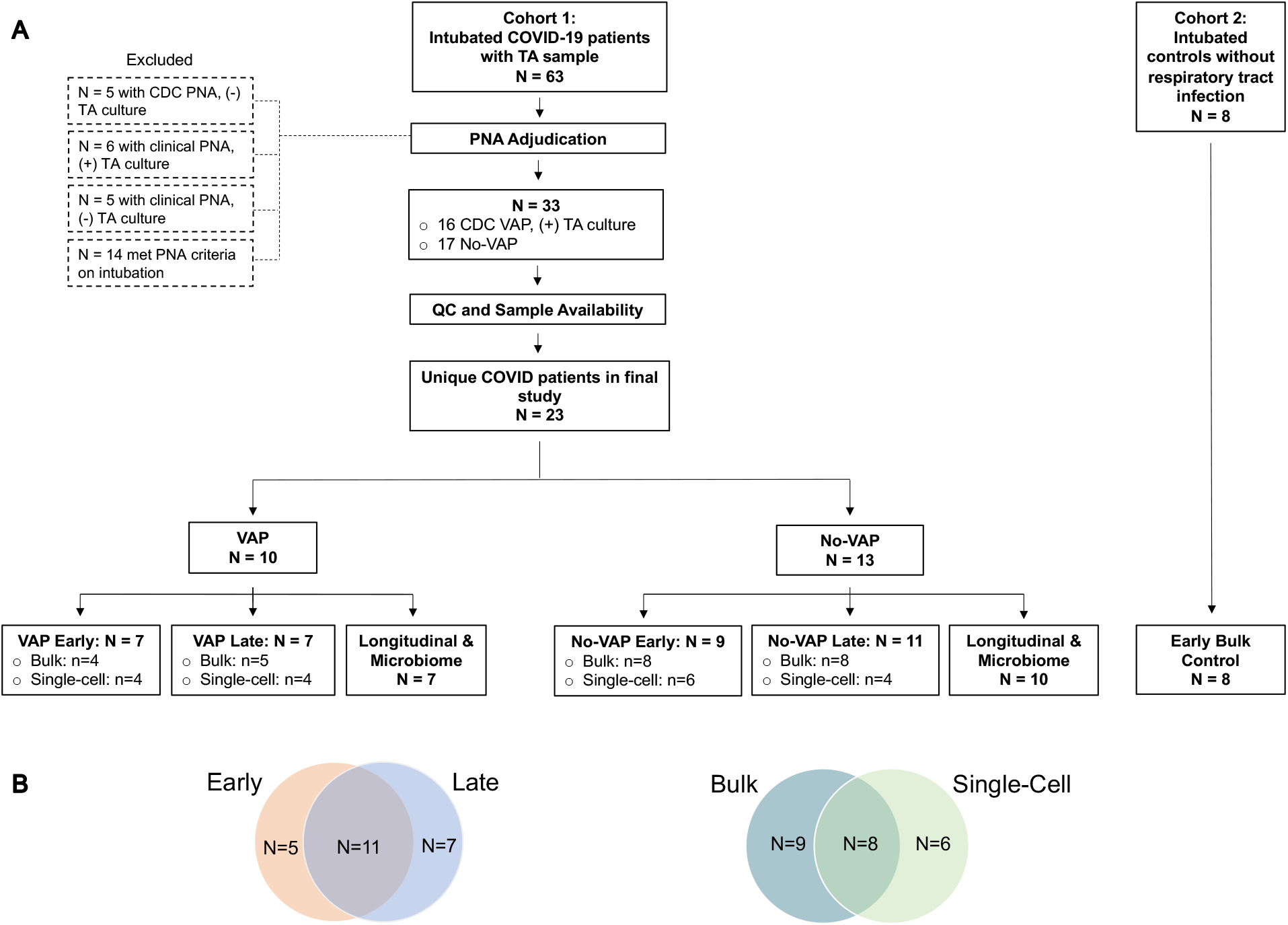
Study flowchart. **A)** Two patient cohorts were studied. Cohort 1 consisted of COVID-19 patients from the COVID Multiphenotyping for Effective Therapies (COMET) / Immunophenotyping Assessment in a COVID-19 Cohort (IMPACC) studies (described in Methods). Cohort 2 consisted of critically ill intubated control patients from a prior prospective cohort study led by our research group^22^. The “early” samples were the first available tracheal aspirate specimens after intubation, and for the VAP group were collected >7 days prior to VAP onset. The “late” samples were obtained 2-4 days before VAP onset for the VAP group; for the No-VAP group, samples were selected to match the VAP group’s time from intubation as closely as possible. Controls included eight critically ill, mechanically ventilated patients without LRTI. All COVID-19 patients included in the primary bulk analysis were also included in the longitudinal host expression and microbiome analyses. Patients who met CDC VAP criteria with negative TA culture, patients who met clinical VAP criteria, and patients with pneumonia upon intubation were excluded. See Supplementary Data File 8 for more details. **B)** The Venn diagram on the left illustrates that patients were included at the “early” timepoint, the “late” timepoint, or both, depending on sample availability. The Venn diagram on the right shows that subjects were included in the bulk sequencing analysis, the single-cell analysis, or both, again dependent on sample availability. Abbreviations: VAP=ventilator-associated pneumonia; TA=tracheal aspirate; QC=quality control; PNA=pneumonia; CDC=United States Centers for Disease Control and Prevention.

Patients with VAP were adjudicated using the United States Centers for Disease Control (CDC) definition^20^, including a requirement for a positive bacterial TA culture (n=10). We defined onset of VAP as the first day a patient developed any of the criteria used to meet the definition, in accordance with CDC guidance^20^. Patients who did not meet the CDC criteria for VAP, and for whom there was no sustained clinical suspicion for bacterial pneumonia during the admission, were adjudicated as No-VAP (n=13). Patients who met CDC VAP criteria but had negative bacterial TA cultures, patients who were clinically suspected of having VAP with either positive or negative TA cultures, and patients who met clinical pneumonia criteria on intubation were excluded from this study **(Figure 1)**.

We compared lower respiratory tract host transcriptional responses between the VAP and No-VAP groups at two timepoints. “Early” timepoint TA samples were collected a median of two days post-intubation and 17 days before VAP onset (range: 15-38 days) for bulk RNA-seq analyses or 10 days before VAP onset (range: 8-16 days) for scRNA-seq analyses. “Late” timepoint samples were collected a median of two days before VAP onset (range: 2-3 days) for bulk RNA-seq analyses and three days before VAP onset (range: 2-4 days) for scRNA-seq analyses, and compared against samples collected from No-VAP patients at similar timepoints post-intubation **(Figure 1, Table 1)**. No samples were analyzed after the onset of VAP. We additionally evaluated eight intubated patients with non-pneumonia illnesses as controls at the “early” timepoint.

**Table 1:** Clinical characteristics of patients with COVID-19 who do or do not develop VAP at the “early” and “late” timepoints. Additional clinical and demographic characteristics can be found in Supplementary Tables 1 (bulk RNA-seq) and 2 (single cell RNA-seq).

|  | VAP (early timepoint) | No-VAP (early timepoint) | P-value (early - VAP vs No-VAP) | VAP (late timepoint) | No-VAP (late timepoint) | P-value (late - VAP vs No-VAP) | Controls (early timepoint) | P-value (+/- COVID-19) |
| --- | --- | --- | --- | --- | --- | --- | --- | --- |
| **Bulk RNA-seq** |  |  |  |  |  |  |  |  |
| N | 4 | 8 |  | 5 | 8 |  | 8 |  |
| Age, median (IQR) | 59.5 (56.0, 66.0) | 49 (40.5, 64.0) | 0.268 | 51.0 (48.0, 53.0) | 59.5 (47.0, 64.5) | 0.271 | 63.0 (36.0, 79.0) | 0.642 |
| Female (%) | 1 (25.0%) | 3 (37.5%) | 1.00 | 2 (40.0%) | 4 (50.0%) | 1.00 | 4 (50.0%) | 0.648 |
| Days before VAP (median [range]) | 16.5 (15, 38) | - | - | 2 (2, 3) | - | - | - | - |
| Days post intubation (median [range]) | 2 (1, 5) | 2 (1, 4) | 0.930 | 17 (8, 23) | 9 (6, 19) | 0.160 | 1 (1, 3) | 0.062 |
| Cultured microbe in VAP patients (n) |  |  |  |  |  |  |  |  |
| *Klebsiella aerogenes* | 2 | - | - | 2 | - | - | - | - |
| *Staphylococcus aureus* | 1 | - | - | 0 | - | - | - | - |
| *Proteus mirabilis* | 1 | - | - | 1 | - | - | - | - |
| *Citrobacter koserii* | 0 | - | - | 1 | - | - | - | - |
| *Serratia marcescens* | 0 | - | - | 1 | - | - | - | - |
| *Pseudomonas aeruginosa* | 0 | - | - | 0 | - | - | - | - |
| **Single cell RNA-seq** |  |  |  |  |  |  |  |  |
| N | 4 | 6 |  | 4 | 4 |  | - | - |
| Age, median (IQR) | 64.0 (50.5, 76.5) | 56.5 (47.0, 68.0) | 0.392 | 61.5 (37.5, 74.0) | 60.0 (50.5, 74.0) | 0.271 | - | - |
| Female (%) | 2 (50.0%) | 2 (33.3%) | 1.00 | 0 (0.0%) | 2 (50.0%) | 0.429 | - | - |
| Days before VAP, median (range) | 9.5 (8, 16) | - | - | 3 (2, 4) | - | - | - | - |
| Days after intubation, median (range) | 5.5 (1, 10) | 4 (2, 13) | 0.748 | 8.5 (7, 18) | 10.5 (4, 15) | 0.884 | - | - |
| Cultured microbe in VAP patients (n) |  |  |  |  |  |  |  | - |
| *Staphylococcus aureus* | 1 | - | - | 0 | - | - | - | - |
| *Klebsiella aerogenes* | 1 | - | - | 1 | - | - | - | - |
| *Citrobacter koserii* | 1 | - | - | 1 | - | - | - | - |
| *Proteus mirabilis* | 0 | - | - | 0 | - | - | - | - |
| *Serratia marcescens* | 0 | - | - | 1 | - | - | - | - |
| *Pseudomonas aeruginosa* | 1 | - | - | 1 | - | - | - | - |

Statistical significance was determined using Fisher's exact test (categorical variables) or by Wilcoxon test (continuous variables). Abbreviations: IQR: interquartile range; VAP: ventilator-associated pneumonia.

We considered that sex as a biologic variable is important for acute COVID-19 severity, and thus asked whether differences in sex might explain our observations. However, no differences in sex were present between VAP and No-VAP groups (P=1, early and late timepoints) **(Table 1)**. There were also no significant differences between groups with respect to age, race or ethnicity **(Table 1, S1, S2)**. In addition, there were no differences between groups with respect to in-hospital receipt of any immunosuppressant or antibiotics prior to sample collection (**Table S1, S2, S3**).

### COVID-19 VAP is associated with a transcriptional signature of bacterial infection two days before VAP onset

We began by assessing the lower respiratory host transcriptional response at the “late” timepoint, just before VAP onset. We carried out differential gene expression analysis on TA bulk RNA-seq data from five patients who developed VAP and eight No-VAP patients **(Table 1)**. We identified 436 differentially expressed genes at a False Discovery Rate (FDR) < 0.1 **(Figure 2A)** and performed gene set enrichment analysis (GSEA) **(Figure 2B)**.

**Figure 2:**
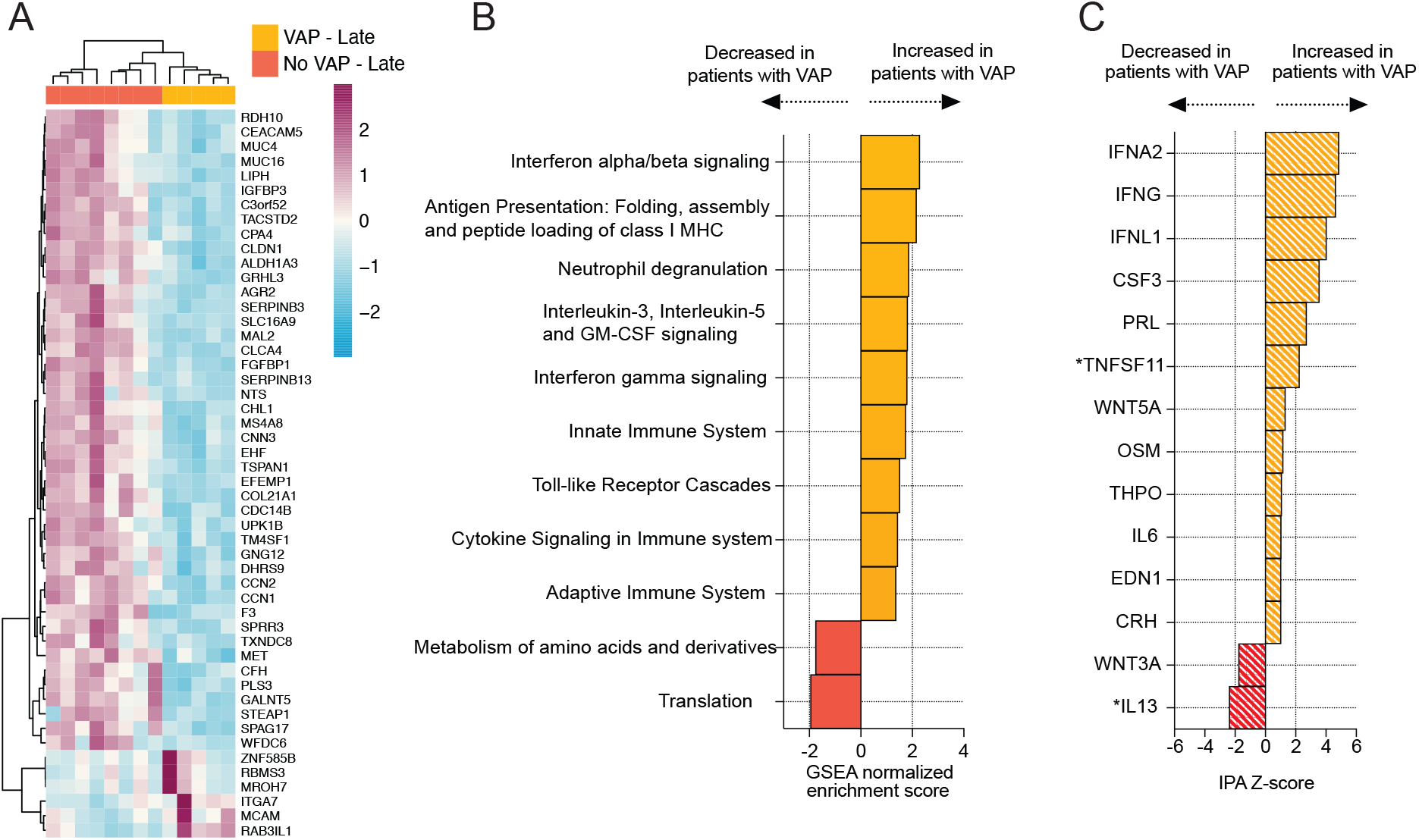
COVID-19 VAP is associated with a lower respiratory tract transcriptional signature of bacterial infection 2 days before VAP onset. **A)** Heatmap of the top 50 differentially expressed genes by adjusted P-value between COVID-19 patients who developed VAP (yellow) versus those who did not (red) at the “late” timepoint, two days before the onset of VAP, from bulk RNA-seq. **B)** Gene set enrichment analysis (GSEA) at the “late” timepoint based on differential gene expression analyses. GSEA results were considered significant with an adjusted P-value < 0.05. **C)** Ingenuity Pathway Analysis (IPA) of upstream cytokines at the “late” timepoint based on differential gene expression analyses. IPA results were considered significant with a Z-score absolute value > 2 and overlap P-value < 0.05. *Denotes cytokines that were included with an overlap P-value < 0.1. All pathways and cytokines are shown in Supplementary Data Files 2 and 3.

As expected, the VAP group exhibited upregulation of pathways related to antibacterial immune responses in the days preceding secondary infection, such as neutrophil degranulation, toll-like receptor signaling, cytokine signaling and antigen presentation **(Figure 2B)**. We also noted that interferon alpha/beta signaling was upregulated. Ingenuity pathway analysis (IPA) predicted broad activation of upstream inflammatory cytokines (including IFN*α* and IFN*γ*) in the VAP group at this “late” timepoint **(Figure 2C)**.

### COVID-19 patients who develop VAP have attenuated immune signaling two weeks before VAP onset

Given our findings of a unique lower respiratory transcriptional signature in the days preceding VAP onset, we next asked whether differences in host immune signaling might exist even earlier, two or more weeks before clinical diagnosis of VAP. We hypothesized that such differences might provide insight regarding the increased susceptibility to secondary bacterial infection in these patients.

We identified 154 differentially expressed genes at the “early” timepoint **(Table 1)**. The patients who developed VAP exhibited lower expression of several genes with established roles in innate immunity including *IFI30, MMP2, TLR9*, and *DEFB124* **(Figure 3A)**. GSEA further revealed that patients who developed VAP had lower expression of pathways related to antibacterial immune responses including neutrophil degranulation, toll-like receptor signaling, IL-17 signaling, antigen presentation and complement pathways, and higher expression of interferon-alpha/beta signaling pathways, more than two weeks before the onset of VAP **(Figure 3B)**. Additionally, pathways related to adaptive immunity such as T and B cell receptor signaling were downregulated in the COVID-19 patients who later developed VAP **(Figure 3B)**.

**Figure 3:**
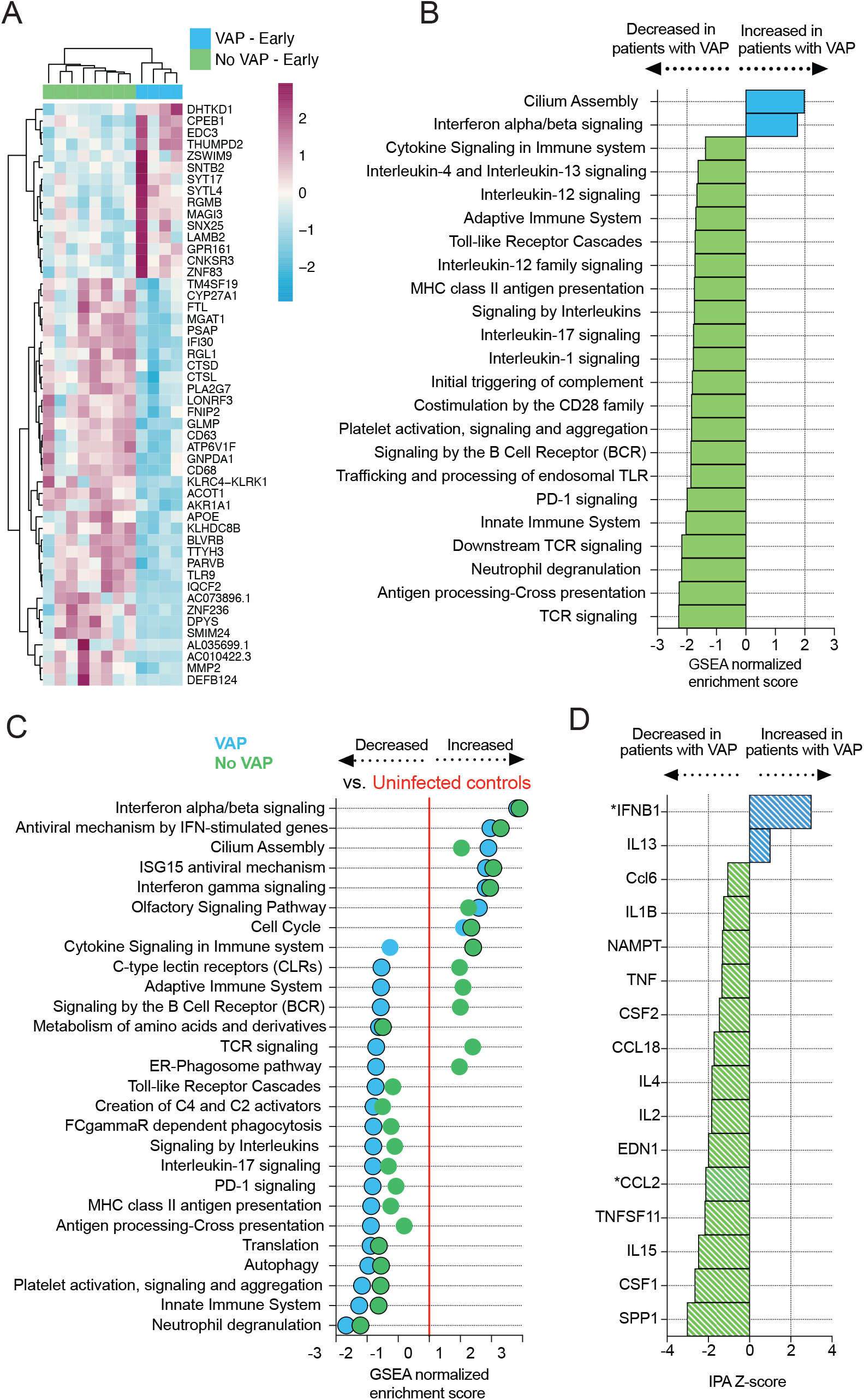
COVID-19 patients who develop VAP have attenuated immune signaling in the lower respiratory tract two weeks before onset of secondary bacterial pneumonia. **A)** Heatmap of the top 50 differentially expressed genes by adjusted P-value between COVID-19 patients who developed VAP (blue) versus those who did not (green) at the “early” timepoint from bulk RNA-seq. **B)** Gene set enrichment analysis at the “early” timepoint based on differential gene expression analyses. GSEA results were considered significant with an adjusted P-value < 0.05. **C)** Expression of GSEA pathways at the “early” timepoint with respect to a baseline of uninfected, intubated controls. Pathways were selected from the GSEA results if they had an adjusted P-value < 0.05 in at least one of the comparisons (VAP vs controls or No-VAP vs controls). Pathways with an adjusted P-value < 0.05 when compared to controls are indicated by circles with a black outline. **D)** Ingenuity Pathway Analysis (IPA) of upstream cytokine activation states at the “early” timepoint based on differential gene expression analyses. IPA results were considered significant with a Z-score absolute value > 2 and overlap P-value < 0.05. *Denotes cytokines that were included with an overlap P-value < 0.1. All pathways and cytokines are shown in Supplementary Data Files 2 and 3.

To gauge the degree of immune pathway suppression compared to controls, we performed a similar analysis on critically ill intubated patients without infection **(Figure 3C)**. Relative to the control patients, multiple antibacterial immune pathways were downregulated in both COVID-19 groups, with the greatest attenuation in the VAP group **(Figure 3C)**. IPA upstream cytokine analysis predicted reduced activation of multiple proinflammatory cytokines juxtaposed against increased activation of IFNB1 in the VAP patients **(Figure 3D)**. Compared to the control group, the predicted activation states of several proinflammatory cytokines were decreased in both COVID-19 groups **(Figure S2)**.

Given prior reports demonstrating a correlation between SARS-CoV-2 viral load and interferon-related gene expression^21^ we next asked whether viral load differed between VAP and No-VAP patients. At the “early” timepoint, no differences were observed in either PCR cycle threshold or bulk RNA-seq SARS-CoV-2 reads per million (P=0.53 and P=0.84, respectively) **(Figure S3)**. We also considered the possibility that differences in the number of days of steroid exposure prior to sample collection might explain our findings but found no differences between groups (P=0.34) **(Table S1, S3)**.

### COVID-19 VAP is associated with impaired antibacterial immune signaling in monocytes and macrophages

To further understand the observed downregulation of key antibacterial signaling pathways in COVID-19 patients who developed VAP, we next asked whether this was driven by specific types of immune cells. We performed scRNA-seq on TA specimens and enriched for immune cells using CD45 selection **(Methods)**. We analyzed 12,197 cells from 14 patients after quality control and batch correction. First, we analyzed samples obtained post-intubation at the “early” timepoint. Clustering based upon cellular transcriptional signatures followed by a comparison of cell type proportions did not reveal statistically significant differences between VAP and No-VAP groups with respect to any cell type at the “early” timepoint **(Figure 4A, S4**). Monocytes and macrophages were the most abundant cell types, and thus we focused transcriptional assessment on these two populations.

**Figure 4:**
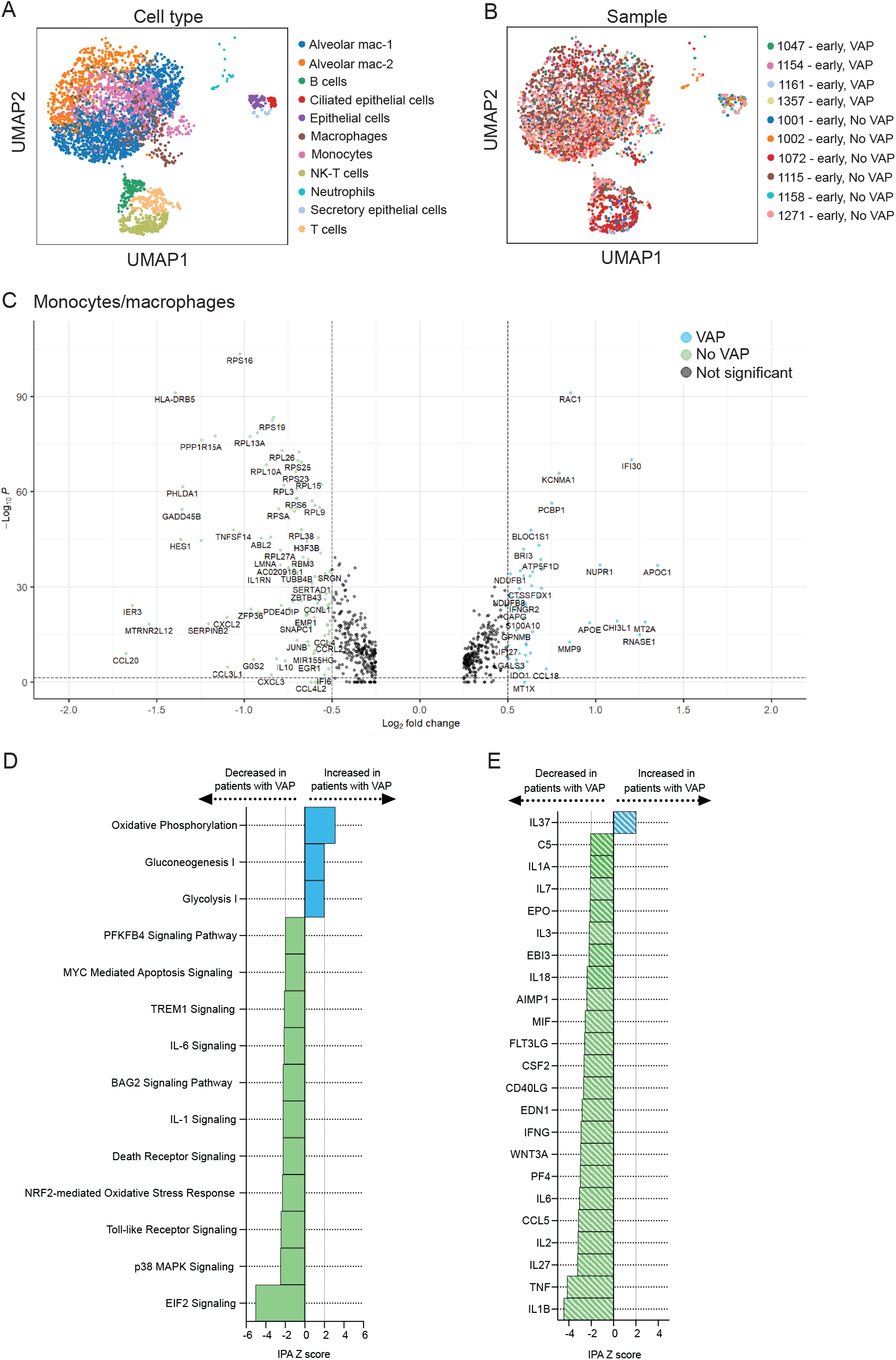
scRNA-seq demonstrates that COVID-19 VAP is associated with early impaired antibacterial immune signaling in lower respiratory tract monocytes and macrophages. **A-B)** UMAP of single cell RNA-seq data from patients that do or do not develop VAP at the “early” timepoint, annotated by (A) cell type or (B) sample. **C)** Volcano plot displaying the differentially expressed genes between VAP and No-VAP patients in the low-resolution monocytes/macrophages cluster. **D-E)** Ingenuity Pathway Analysis (IPA) of (D) key canonical pathways and (E) upstream cytokines based on differential gene expression analysis in monocytes/macrophages of COVID-19 patients who developed VAP versus those who did not. Only significant pathways (IPA Z-score absolute value > 2 and overlap P-value < 0.05) are depicted. All pathways and cytokines are provided in Supplementary Data Files 5 and 6.

COVID-19 patients who developed VAP had distinct post-intubation transcriptional signatures compared to the No-VAP patients at this “early” timepoint, with respect to the overall monocyte/macrophage cluster **(Figure 4, S4, S5)**. We identified 625 differentially expressed genes (FDR < 0.05) in this cluster and noted downregulation of several genes with key roles in innate immunity in the COVID-19 patients who subsequently developed VAP compared to those who did not, including *IL1Rn, CXCL2, CCL20*, and *JunB*. (Figure 4C). In addition, we noted upregulation of interferon-induced genes including *IFI30* and *IFNγR-2*. **(Figure 4C)**. Findings were similar when evaluating differential expression within alveolar macrophages, the largest monocyte/macrophage subset (**Figure S5**).

IPA canonical pathway analysis revealed downregulation of several cytokine and innate immune signaling pathways in the patients who later developed VAP at the “early” post-intubation timepoint, including IL-6 and IL-1 signaling, and the NRF2-mediated oxidative stress response **(Figure 4D)**. Computational analysis of upstream cytokine activation predicted impaired activation of multiple pro-inflammatory cytokines in patients who developed VAP, including TNF, IFNγ, and IL1B **(Figure 4E)**. Similar findings were observed in the alveolar macrophage population **(Figure S5)**. We were unable to reliably assess the T cell population due to low cell numbers.

A comparison of monocytes and macrophages between the VAP and No-VAP patients just prior to VAP onset at the “late” timepoint revealed significant upregulation of genes essential for antibacterial defense. These findings reflected bulk RNA-seq observations and were characterized by upregulation in the VAP group of pathways related to IL-17 and nitric oxide signaling, reactive oxygen species production, and hypercytokinemia **(Figure S6)**.

### Temporal dynamics of the host response in COVID-19 patients who develop VAP

We next investigated temporal dynamics of the lower airway host immune response in COVID-19 patients from the time of intubation to development of VAP, by evaluating bulk RNA-seq differential gene expression between COVID-19 VAP patients at the “early” timepoint (median of 17 days before VAP onset, n=4) versus “late” timepoint (median of two days before VAP onset, n=5). We identified 2705 differentially expressed genes, and unsupervised hierarchical clustering of the 50 most significant genes demonstrated clear separation of the two timepoints **(Figure 5A)**. GSEA revealed that type I interferon signaling was notably downregulated at the “late” timepoint in comparison to the “early” timepoint **(Figure 5B)**; however, expression of this pathway was still significantly higher in the VAP versus the No-VAP patients **(Figure 2B)**. Several other immune signaling pathways were more highly expressed at the “late” timepoint, presumably reflecting activation of an antibacterial response in the setting of impending bacterial pneumonia **(Figure 5B)**. Consistent with this, upstream regulator analysis predicted increased activation of several pro-inflammatory cytokines and decreased IFN*α* and IFN*λ* signaling at the “late” versus “early” timepoints **(Figure 5C)**.

**Figure 5:**
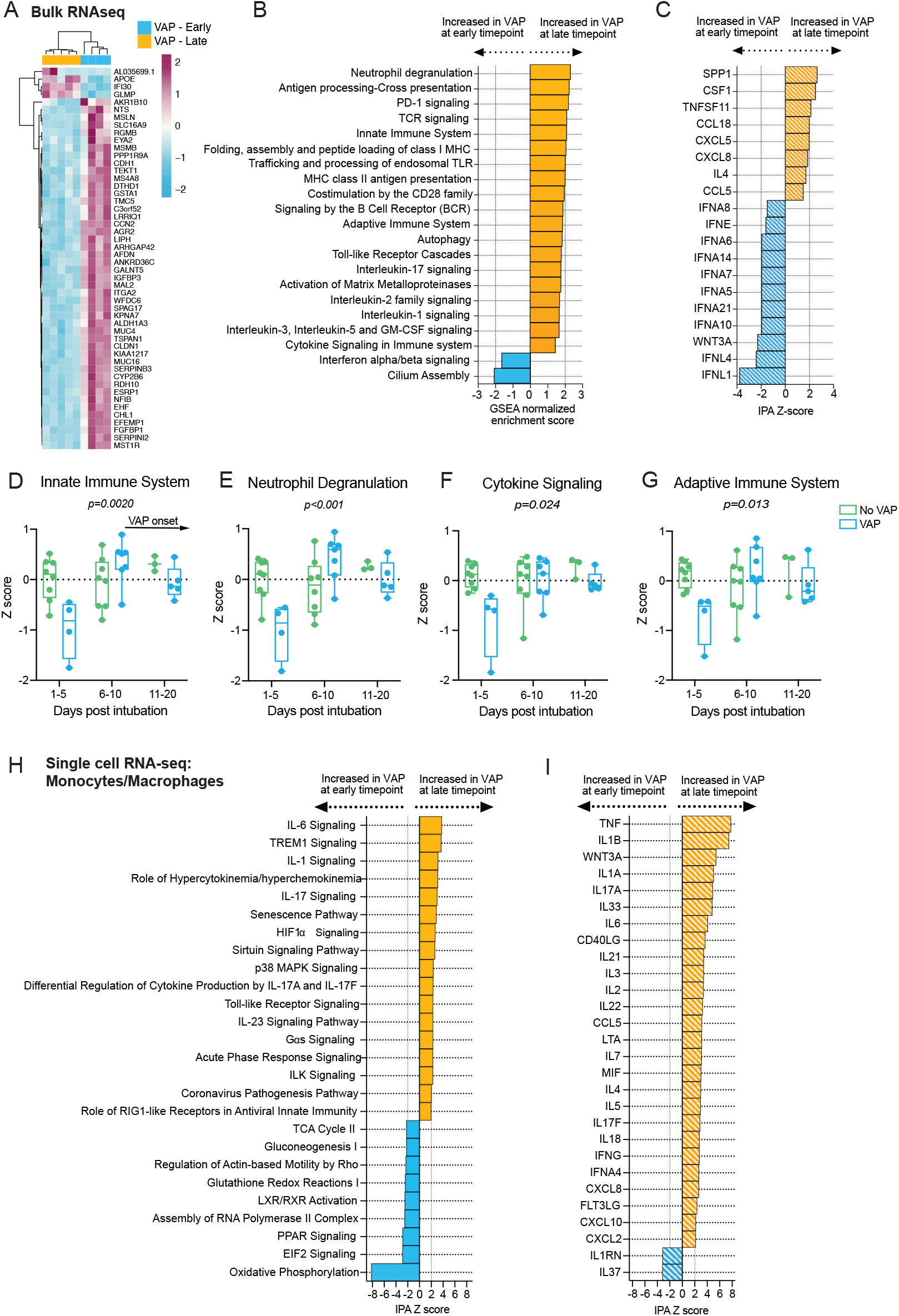
Temporal dynamics of the host response to VAP. **A)** Heatmap of the top 50 differentially expressed genes by adjusted P-value between COVID-19 patients who developed VAP at the “early” timepoint (blue) versus the “late” timepoint (yellow) from bulk RNA-seq. **B)** Gene set enrichment analysis (GSEA) based on differential gene expression of VAP patients at the “early” vs “late” timepoint from bulk RNA-seq. GSEA results were considered significant with an adjusted P-value < 0.05. **C)** Ingenuity Pathway Analysis (IPA) of upstream cytokine activation states based on differential gene expression analyses of VAP patients at the “early” vs “late” timepoint from bulk RNA-seq. IPA results were considered significant with a Z-score absolute value > 2 and overlap P-value < 0.05. **D-G)** Longitudinal analysis of selected pathway expression in VAP (blue) versus No-VAP (green) patients from bulk RNA-seq samples taken from time of intubation to onset of VAP for all patients. Pathway Z-scores were calculated by averaging Z-scores for the top 20 leading edge genes of each pathway, determined by the results of GSEA comparing VAP versus No-VAP patients at the “early” timepoint. Multiple Z-scores per patient at a given time interval were averaged so that each patient corresponds to one datapoint at each interval. Samples from day 21+ after intubation are not depicted due to a lack of these later timepoints in the No-VAP group. VAP onset in these patients ranged from 10-39 days post intubation (median: 19 days). Selected pathways are: innate immune system (D), neutrophil degranulation (E), cytokine signaling (F), and adaptive immune system (G). Box plots represent the median and range. Statistical significance was determined by two-way ANOVA, and interaction P-values are highlighted. **H-I)** Ingenuity Pathway Analysis (IPA) of (H) key canonical pathways (I) and upstream cytokine activation states based on differential gene expression analysis in monocytes/macrophages from scRNA-seq of COVID-19 patients who developed VAP versus those who did not. Only significant pathways (IPA Z-score absolute value > 2 and overlap P-value < 0.05) are depicted. All pathways and cytokines are provided in Supplementary Data Files 2, 3, 5, and 6.

In contrast, comparing No-VAP patients at the “early” (n=8) versus “late” (n=8) timepoints yielded only two genes, both of which were interferon-stimulated genes (*RSAD2* and *CMPK2*) downregulated at the “late” timepoint, suggesting that while the host response was relatively unchanged in these patients, the antiviral response attenuated over time. Indeed, GSEA revealed that type I interferon signaling, and other antiviral immune pathways, were downregulated in the patients who did not develop VAP at the “late” timepoint **(Figure S7)**.

We further compared dynamics of host immune responses between VAP and No-VAP patients by performing longitudinal analyses of key immune signaling pathways, including all patients with available bulk RNA-seq samples (VAP: n=7, No-VAP: n=10). Onset of VAP ranged from 10-39 days post intubation (median 19 days), and treatment with immunosuppressants did not differ significantly between comparator groups (p=0.30, Fisher’s exact test). Early attenuation of immune signaling in the VAP group was conspicuous, and this pattern eventually resolved later in disease course by the time secondary bacterial infection became established **(Figures 5D-G)**. We confirmed that the observed differences between VAP and No-VAP patients were not driven by differences in treatment with immunosuppressants by comparing pathway Z-scores between patients who received immunosuppressants and those who did not, at the “early” timepoint **(Figure S8)**.

We subsequently performed a similar comparison between the “early” and “late” timepoints based on scRNA-seq data from patients who developed VAP. Differential gene expression analysis between these two populations identified 1466 differentially expressed genes in the encompassing monocyte/macrophage cluster. IPA on these genes revealed upregulation of antibacterial signaling pathways at the “late” timepoint, including signaling by several cytokines (including IL-17, IL-6, TNF, IFN) **(Figure 5H-I)**, congruent with the bulk RNA-seq analysis.

### Lung microbiome disruption precedes VAP in COVID-19 patients

We hypothesized that the innate immune suppression observed in patients who developed VAP would correlate with SARS-CoV-2 viral load. Using TA metatranscriptomics to assess the lower respiratory microbiome, we evaluated longitudinal changes in SARS-CoV-2 abundance. Although no difference in viral load was observed at the “early” timepoint **(Figure S3, Figure 6A)**, the trajectory of SARS-CoV-2 viral load differed significantly in patients who developed VAP. Viral load decreased over time in both groups, however the virus was still detected at later timepoints in patients with VAP, while it was undetected in any of the No-VAP patients at these timepoints (Figure 6A). This result suggested that COVID-19 patients who develop VAP may exhibit impaired ability to clear the virus compared to those who do not, and that the lung microbiome composition may be similarly impacted.

**Figure 6:**
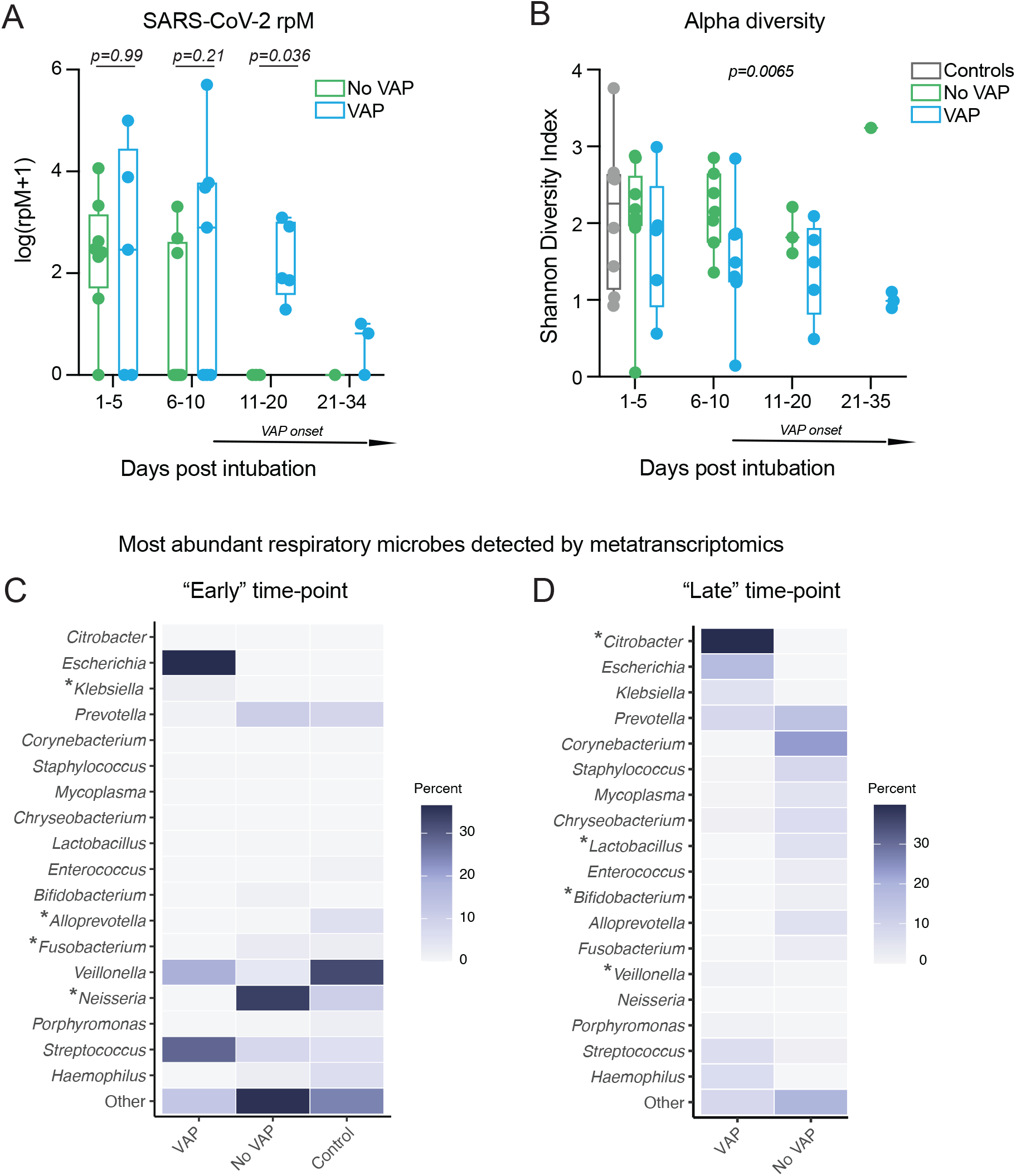
Lung microbiome community collapse precedes VAP in COVID-19 patients. **(A)** SARS-CoV-2 viral load (reads per million sequenced, rpM) over time by days since intubation in patients who develop VAP vs those who do not. For plotting purposes, log(rpM+1) was used to avoid negative values. **(B)** Lung microbiome bacterial diversity (Shannon’s Index) in COVID-19 patients with relation to VAP development over time by days since intubation. VAP onset in these patients ranged from 10-39 days post intubation (median: 19 days). SARS-CoV-2 negative intubated controls are included for comparison. Box plots represent the median and range. Statistical significance was determined by Mann-Whitney tests (A) or two-way ANOVA between VAP and No-VAP patients (B). P-values < 0.05 were considered significant. **(C-D)** Most abundant respiratory microbes detected by metatranscriptomics at the (C) “early” and (D) “late” timepoints. The mean abundance of the top 10 most abundant microbes from each group is depicted, and significantly enriched taxa in VAP patients compared to No-VAP patients are marked by an asterisk. SARS-CoV-2 negative intubated controls are included for comparison at the “early” timepoint. Microbe abundance data for each patient can be found in Supplementary Data File 7.

Indeed, COVID-19 patients who developed VAP exhibited a significant reduction in bacterial alpha diversity of their airway microbiome before clinical signs of infection (Shannon Diversity Index, P=0.0065; Figure 6B). Beta diversity did not differ significantly, which may be due to the fact that VAP was caused by different pathogens in each patient, resulting in different airway microbiome compositions in each case **(Figure S9)**. All patients received antibiotics prior to collection of the first sample, suggesting that antibiotic use was not driving these differences **(Table S1)**.

We next assessed the most abundant bacterial genera in the lung, as identified by TA metatranscriptomics. At the “early” timepoint, control and No-VAP samples were predominated by common lung commensals, with significant enrichment (FDR < 0.05) of *Alloprevotella*, *Fusobacterium*, and *Neisseria* compared to the VAP group (Figure 6C). In the VAP group, early significant enrichment of *Klebsiella* was noted, suggestive of a shift in the microbiome structure that precedes disease onset and culture positive confirmation (Figure 6C). By the “late” timepoint, VAP-associated pathogens such as *Citrobacter*, *Escherichia*, and *Klebsiella* were highly prevalent. *Citrobacter* was significantly enriched in VAP versus No-VAP groups at this timepoint, supporting culture results. While highly enriched in VAP, *Klebsiella* did not reach significance at this timepoint due to high variance between individuals (Figure 6D). No-VAP patients had a significantly greater abundance of commensal lung microbes, such as *Lactobacillus*, *Bifidobacterium*, and *Veillonella,* at the “late” timepoint **(Figure 6D)**.

## Discussion

Secondary bacterial pneumonia contributes to significant morbidity and mortality in patients with primary viral lower respiratory tract infections^1, 3^, but mechanisms governing individual susceptibility to VAP have remained unclear. Few human cohort studies have evaluated the immunologic underpinnings of VAP, and none have been reported in the context of COVID-19, which is characterized by a dysregulated host response distinct from other viral pneumonias^21–23^. To address this gap and probe mechanisms of VAP susceptibility in patients with COVID-19, we carried out a systems biological assessment of host and microbial dynamics of the lower respiratory tract.

Two days before VAP onset, a transcriptional signature consistent with bacterial infection was observed. This finding suggests that host response changes can occur before clinical recognition of pneumonia, highlighting the potential utility of the host transcriptome as a tool for VAP surveillance. While intriguing, this observation did not provide an explanation for differential susceptibility of some COVID-19 patients to post-viral pneumonia.

The discovery of an early suppressed antibacterial immune response in patients who later developed VAP did, however, offer a potential explanation. More than two weeks before VAP onset, we observed a striking suppression of pathways related to both innate and adaptive immunity, including neutrophil degranulation, toll-like receptor (TLR) signaling, complement activation, antigen presentation, and T cell receptor and B receptor signaling, as well as signaling by several cytokines (e.g. IL-1, IL-4, IL-12, IL-13 and IL-17). Comparison against uninfected, intubated controls confirmed the previously described paradoxical impairment in immune signaling found in patients with severe COVID-19^22^, and suggested that VAP susceptibility may be the result of disproportionate suppression of innate and adaptive pathways critical for antibacterial defense, resulting in enhanced susceptibility to opportunistic secondary infections.

Animal models of influenza have provided insight into potential mechanisms of post-viral pneumonia, although none have provided insight regarding why some individuals are more susceptible than others. In mice inoculated with influenza, for instance, virus-induced type I interferon suppresses neutrophil chemokines and impairs Th17 immunity, compromising effective clearance of bacterial infections^10, 11^. Interestingly, we also observed increased type I interferon signaling in COVID-19 patients who weeks later developed VAP, and an impairment in IL-17 signaling and other immune pathways. Desensitization to TLR ligands after influenza infection has also been documented^24^, which is congruent with the downregulation of TLR signaling at the time of intubation observed in our bulk RNA-seq analyses.

We asked whether certain cell types were responsible for driving the early suppression of immune signaling observed in COVID-19 patients who went on to develop VAP. No significant differences in proportions of any cell types was observed between patients with or without VAP at the time of intubation. This finding suggests that an impairment of immune cell recruitment was not causing these differences, but rather significant gene expression differences within these immune cell populations.

In the most predominant cell population, the monocytes and macrophages, we observed broad downregulation of the innate and adaptive immune responses, concordant with global observations in bulk RNA-seq analyses. Further, we noted a downregulation of key pathways involved in antimicrobial immune responses including TREM1, IL-1, and IL-6 signaling. Overall, our data suggest that while no difference in cell type populations existed between groups, changes in the gene expression within monocytes and macrophages contributes to immune suppression in COVID-19 patients who later develop VAP.

SARS-CoV-2 viral load correlates with interferon stimulated gene expression^21, 22^ and thus we initially hypothesized that differences in viral load between groups might relate to individual VAP susceptibility. However, we found no difference between groups at the “early” timepoint. Moreover, no differences existed in terms of immunosuppressive medication administration or clinically diagnosed immunodeficiency, suggesting that other, still unidentified mechanisms present at the time of intubation must underlie the marked suppression of immune gene expression in COVID-19 patients who went on to develop VAP.

While no difference in viral load was observed at the time of intubation, the COVID-19 patients who developed VAP exhibited impaired viral clearance over the time-course of intubation. This observation was corroborated by a prolonged antiviral type I interferon response at the “late” timepoint (median of two days before VAP onset) in patients who developed VAP versus those who did not, pointing to the persistence of suboptimal antiviral immunity in these patients. Early induction of functional SARS-CoV-2 specific T cells is associated with faster viral clearance in COVID-19 patients^25^ and likewise, we observed impairments in T cell activation and signaling in the VAP group by bulk RNA-seq at the “early” timepoint, which further suggests a decreased ability to control the virus in these patients.

Respiratory viruses can reshape the human airway microbiome by modulating host inflammatory responses^26, 27^. In mouse models of influenza, the airway microbiome exhibits expansion of several bacterial families during the course of viral infection as innate immunity is suppressed^26^. These changes increase the risk of secondary bacterial infection^26^ and have been observed in patients with chronic obstructive pulmonary disease, where suppression of the innate immune response in rhinovirus infected patients may be followed by bacterial superinfection^28, 29^. Similarly, the innate immune suppression observed in COVID-19 patients who developed

VAP was associated with airway microbiome collapse and the outgrowth of lung pathogens in advance of clinical VAP diagnosis. This finding suggests that individual immune responses to SARS-CoV-2 infection may drive a restructuring of the microbial community and increase susceptibility to VAP (Figure 7). The resulting outgrowth of a VAP-associated bacterial pathogen may elicit an antibacterial response, but the broader immunosuppressive state preceding this response may be insufficient to control the development of clinical pneumonia. Those with a lesser degree of immunosuppression may be able to respond faster and therefore control opportunistic bacterial pathogens more effectively.

**Figure 7:**
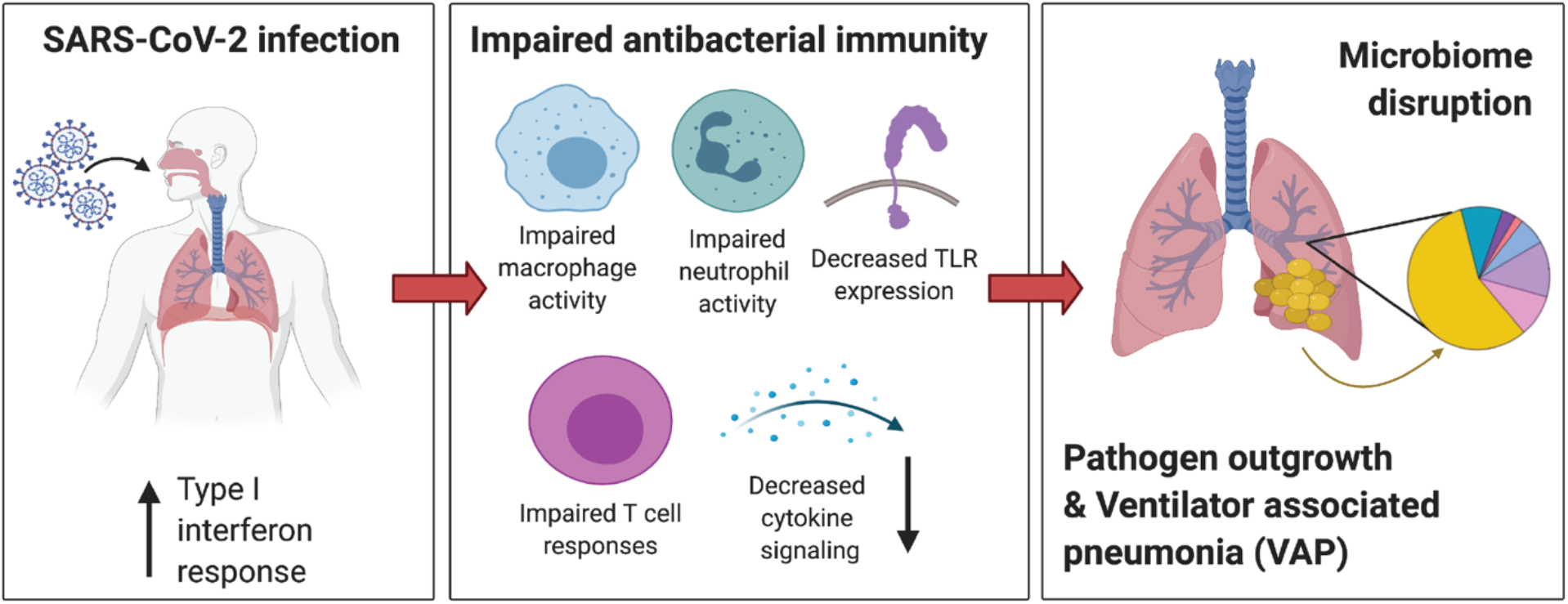
Mechanistic hypothesis of secondary bacterial pneumonia susceptibility in patients with COVID-19. We hypothesize that individual immune responses to SARS-CoV-2 infection may drive a restructuring of the microbial community and increase susceptibility to VAP. Those predisposed to VAP have increased type I interferon responses and dysregulated antibacterial immune signaling characterized by impaired macrophage, neutrophil and T cell activity, decreased toll-like receptor (TLR) signaling and impaired activation of cytokines important for pathogen defense. This state of suppressed immunity may disrupt the lower respiratory tract microbiome, predisposing certain patients to the outgrowth of bacterial pathogens and VAP. Created with BioRender.com.

These findings may also have important implications for management of patients with COVID-19 related acute respiratory failure, many of whom are now being treated with corticosteroids and other immunosuppressive drugs. These agents may lead to further suppression of the key pathways required for host response to secondary bacterial infection. Thus, our results emphasize the need for ongoing vigilance for VAP in patients treated with potent immunosuppressive agents, as well as the need to develop novel diagnostic and/or prognostic approaches to identifying patients at highest risk.

Protein biomarkers of immune function have been associated with nosocomial infection in critically ill patients^15^, although not in the context of viral infection. Further studies with a larger cohort may allow for the identification of transcriptomic biomarkers to assess a COVID-19 patient’s risk of VAP at the time of intubation, which could reduce inappropriate use of prophylactic antibiotics or immunomodulatory treatments, or signal a need for enhanced surveillance strategies.

Sample size is a limitation of this study, which could result in over-fitting to this small set of patients; however, the reproducibility of our observations across both bulk and scRNA-seq analyses and the significant number of differentially expressed genes among the comparator groups support the internal validity of our conclusions. Additional studies in a larger independent cohort will be needed to determine the utility of lower respiratory gene expression biomarkers for predicting future development of VAP. Because this study was limited to critically ill, intubated patients, we were unable to assess early stages of COVID-19, which may provide additional insight regarding determinants of secondary bacterial infection.

We were unable to assess whether epithelial cells contributed to VAP risk due to enrichment for immune cells prior to scRNA-seq. However, given that monocytes, macrophages and lymphocytes can both be infected by SARS-CoV-2 and contribute to disease pathophysiology and resolution^30–32^, our findings are apropos. With larger cohorts, the early detection of specific immune pathway suppression and microbiome collapse could be leveraged to develop clinically useful models for identifying COVID-19 patients with increased susceptibility to secondary bacterial pneumonia.

## Materials and Methods

### Study design, cohorts, and enrollment

We conducted a prospective case-control study of adults requiring mechanical ventilation for COVID-19 with or without secondary bacterial pneumonia. We also evaluated control patients requiring mechanical ventilation for other reasons who had no evidence of pulmonary infection (Figure 1). Patients were enrolled in either of two prospective cohort studies of critically ill patients at the University of California, San Francisco (UCSF) and Zuckerberg San Francisco General Hospital between 07/2013 and 07/2020^33^. Of the COVID-19 patients, 17 were co-enrolled in the National Institute of Allergy and Infectious Diseases-funded Immunophenotyping Assessment in a COVID-19 Cohort (IMPACC) Network study.

### Ventilator-associated pneumonia adjudication

A total of 63 adults who required intubation for severe COVID-19 (Cohort 1) and who had available TA samples were considered for inclusion in the study (Figure 1). Patients who met the US Centers for Disease Control (CDC) definition for VAP^20^ (based on the PNU1 and PNU2 definitions) and had a positive bacterial respiratory culture were adjudicated as having VAP for the purpose of the study (n=16). Patients who did not meet these criteria, and for whom there was no sustained clinical suspicion for bacterial pneumonia during the admission, were categorized as No-VAP (n=17).

VAP and No-VAP patients for whom samples at the timepoints of interest were available were included in the analyses (VAP: n=10; No-VAP: n=13). Patients who met CDC-VAP criteria but had negative TA cultures (n=5 patients), as well as patients with clinically suspected bacterial pneumonia who did not meet CDC VAP criteria and had a positive (n=6) or negative (n=5) TA culture were excluded. Fourteen patients who met clinical pneumonia criteria on intubation were also excluded. Eight intubated patients from a recent study^22^ (Cohort 2) were included as controls and were selected because they had previously been adjudicated as having no evidence of lower respiratory tract infection. This group included four patients with acute respiratory distress syndrome (ARDS) due to non-infectious etiologies, and four patients without ARDS who were intubated for other reasons (subdural hematoma (n=1), retroperitoneal hemorrhage (n=1), or neurosurgical procedures (n=2)).

### Tracheal aspirate sampling

Following enrollment, tracheal aspirate (TA) was collected (periodically following intubation for Study 1, or once within three days of intubation for Study 2), without addition of saline wash, and either a) mixed 1:1 with DNA/RNA shield (Zymo Research) for bulk RNA-seq or b) immediately processed in a biosafety level 3 laboratory (BSL3) for scRNA-seq analysis.

### Bulk RNA sequencing and host transcriptome analysis

#### RNA sequencing

To evaluate host and microbial gene expression, metatranscriptomic next generation RNA sequencing (RNA-seq) was performed on TA specimens. Following RNA extraction (Zymo Pathogen Magbead Kit) and DNase treatment, human cytosolic and mitochondrial ribosomal RNA was depleted using FastSelect (Qiagen). To control for background contamination, we included negative controls (water and HeLa cell RNA) as well as positive controls (spike-in RNA standards from the External RNA Controls Consortium (ERCC))^34^. RNA was then fragmented and underwent library preparation using the NEBNext Ultra II RNA-seq Kit (New England Biolabs). Libraries underwent 146 nucleotide paired-end Illumina sequencing on an Illumina Novaseq 6000.

#### Host differential expression

Following demultiplexing, sequencing reads were pseudo-aligned with kallisto^35^ to an index consisting of all transcripts associated with human protein coding genes (ENSEMBL v. 99), cytosolic and mitochondrial ribosomal RNA sequences and the sequences of ERCC RNA standards. Gene-level counts were generated from the transcript-level abundance estimates using the R package tximport^36^, with the scaledTPM method. Samples retained in the dataset had a total of at least 1,000,000 estimated counts associated with transcripts of protein coding genes. Genes were retained for differential expression analysis if they had counts in at least 30% of samples. Differential expression analysis was performed using the R package DESeq2^37^. We modeled the expression of individual genes using the design formula ∼VAPgroup, where VAP groups were “VAP-early”, “No-VAP-early”, “VAP-late” and “No-VAP-late” and used the results() function to extract a specific contrast. Separate comparisons to the control group were performed using the design formula ∼COVID-19-status to compare positive and negative patients.

Significant genes were identified using a Benjamini-Hochberg false discovery rate (FDR) < 0.1. We generated heatmaps of the top 50 differentially expressed genes by FDR. For visualization, gene expression was normalized using the regularized log transformation, centered, and scaled prior to clustering. Heatmaps were generated using the *pheatmap* package. Columns were clustered using Euclidean distance and rows were clustered using Pearson correlation. Differential expression analysis results are provided in (**Supplementary Data File 1**).

#### Pathway analysis

Gene set enrichment analyses (GSEA) were performed using the fgseaMultilevel function in the R package fgsea^38^ and REACTOME^39^ pathways with a minimum size of 10 genes and a maximum size of 1,500 genes. All genes were included in the comparison, pre-ranked by the test statistic. Significant pathways were defined as those with a Benjamini-Hochberg adjusted P-value < 0.05. Ingenuity Pathway Analysis (IPA) Canonical Pathway and Upstream Regulator Analysis^40^ was employed on genes with P < 0.1 and ranked by the test statistic to identify cytokine regulators. Significant IPA results were defined as those with a Z-score absolute value greater than 2 and an overlap P-value < 0.05. The gene sets in figures were selected to reduce redundancy and highlight diverse biological functions. Full GSEA and IPA results are provided in (**Supplementary Data Files 2 and 3**).

Longitudinal pathway analysis was performed using all available TA samples spanning post-intubation to VAP onset for all patients included in the bulk RNA-seq analysis. Analysis was restricted to samples with at least 1,000,000 human protein coding transcripts. Pathways of interest were selected from the significant GSEA results of the comparison of VAP vs. No-VAP patients in the “early” timepoint. The top 20 leading edge genes were selected from each pathway for analysis. To calculate a Z-score for each gene, expression was normalized using the variance stabilizing transformation (VST), centered, and scaled. A pathway Z-score was calculated by averaging the 20 gene Z-scores. Multiple Z-scores per patient at a given time interval were averaged so that each patient corresponds to one datapoint at each interval. Statistical significance of pathway expression over time between VAP and No-VAP groups was calculated using a two-way analysis of variance (ANOVA) in GraphPad PRISM.

### Single cell RNA sequencing and transcriptome analysis

After collection, fresh TA was transported to a BSL-3 laboratory at ambient temperature to improve neutrophil survival. 3mL of TA was dissociated in 40mL of PBS with 50ug/mL collagenase type 4 (Worthington) and 0.56 ku/mL of Dnase I (Worthington) for 10 minutes at room temperature, followed by passage through a 70μM filter. Cells were pelleted at 350g 4C for 10 minutes, resuspended in PBS with 2mM EDTA and 0.5% BSA, and manually counted on a hemocytometer. Cells were stained with MojoSort Human CD45 and purified by the manufacturer’s protocol (Biolegend). After CD45 positive selection, cells were manually counted with trypan blue on a hemocytometer. Using a V(D)J v1.1 kit according to the manufacturer’s protocol, samples were loaded on a 10X Genomics Chip A without multiplexing, aiming to capture 10,000 cells (10X Genomics). Libraries underwent paired end 150 base pair sequencing on an Illumina NovaSeq6000.

Raw sequencing reads were aligned to GRCh38 using the STAR aligner . Cell barcodes were then determined based upon UMI count distribution. Read count matrices were generated through the 10X genomics cellranger pipeline v3.0. Data was processed and analyzed using the Scanpy v1.6^42^. Cells that had <200 genes, <20% mitochondrial reads, and had greater than 2500 counts were filtered. Mitochondrial genes were removed and multi-sample integration was performed using single-cell variational inference (scVI)^43^. Differential expression was performed using MAST v1.16.0^44^. Differential expression analysis results are detailed in (**Supplementary Data File 4**).

#### Pathway analysis

Ingenuity Pathway Analysis (IPA) Canonical Pathway and Upstream Regulator Analysis^40^ was employed on genes with P < 0.05 and ranked by log2foldchange to identify canonical pathways and cytokine regulators. Due to the significantly greater number of differentially expressed genes in scRNA-seq analyses, we utilized a more restrictive cutoff of FDR < 0.05 for scRNA-seq to ensure a similar number of genes were input into IPA. Significant IPA results were defined as those with a Z-score absolute value greater than 2 and an overlap P-value < 0.05. The gene sets in figures were selected to reduce redundancy and highlight diverse biological functions. Full GSEA and IPA results are provided in (**Supplementary Data Files 5 and 6**).

### Lung microbiome analysis

RNA from tracheal aspirates was sequenced as described above. Taxonomic alignments were obtained from raw sequencing reads using the IDseq pipeline^45, 46^, which performs quality filtration and removal of human reads followed by reference-based taxonomic alignment at both the nucleotide and amino acid level against sequences in the National Center for Biotechnology Information (NCBI) nucleotide (NT) and non-redundant (NR) databases, followed by assembly of reads matching each taxon detected. Taxonomic alignments underwent background correction for environmental contaminants (see below), viruses were excluded, and data was then aggregated to the genus level before calculating diversity metrics. Alpha diversity (Shannon’s

Diversity Index) and beta diversity (Bray-Curtis dissimilarity) were calculated and the latter plotted using non-metric multidimensional scaling (NDMS). Comparison of alpha and beta diversity over time between VAP and No-VAP groups was calculated using a two-way analysis of variance (ANOVA) in R. Top ten taxa in each group were identified by ranking the percent of counts for each genus among total bacterial counts in each group. Significantly enriched taxa were evaluated by comparing distributions using three models (Poisson, negative binomial, and zero-inflated negative binomial mixed-effects models) and correcting for multiple hypothesis testing using Benjamini-Hochberg as previously described^47^. Significantly enriched taxa were called using a q < 0.05 cutoff.

#### Identification and mitigation of environmental contaminants

To minimize inaccurate taxonomic assignments due to environmental and reagent derived contaminants, non-templated “water only” and HeLa cell RNA controls were processed with each group of samples that underwent nucleic acid extraction. These were included, as well as positive control clinical samples, with each sequencing run. Negative control samples enabled estimation of the number of background reads expected for each taxon. A previously developed negative binomial model^21^ was employed to identify taxa with NT sequencing alignments present at an abundance significantly greater compared to negative water controls. This was done by modeling the number of background reads as a negative binomial distribution, with mean and dispersion fitted on the negative controls. For each batch (sequencing run) and taxon, we estimated the mean parameter of the negative binomial by averaging the read counts across all negative controls, slightly regularizing this estimate by including the global average (across all batches) as an additional sample. We estimated a single dispersion parameter across all taxa and batches, using the functions glm.nb() and theta.md() from the R package MASS^48^. Taxa that achieved a P-value < 0.01 were carried forward.

### Statistics

Statistical significance was defined as P < 0.05, using two-tailed tests. Categorical data were analyzed by Fisher’s exact test and nonparametric continuous variables were analyzed by Mann-Whitney. Statistical approaches used for gene expression and microbiome analyses are detailed in each respective Methods section.

### Study approval

Both cohort studies were approved by the UCSF Institutional Review Board (IRB) under protocols 10-02701 (control patients, pre-COVID-19 pandemic) and 20-30497 (COVID-19 patients, COVID-19 Multiphenotyping for Effective Therapy (COMET) study), respectively. Detailed enrollment and consent protocols for both studies have been previously described^18, 33^.

## Supporting information

Data File S1: Differentially expressed genes from bulk RNA-seq analyses.

Data File S2: Gene set enrichment analysis results from bulk RNA-seq analyses. NES = Normalized enrichment score.

Data File S3: Ingenuity pathway analysis for upstream regulating cytokines from bulk RNA-seq analyses.

Data File S4: Differentially expressed genes from single cell RNA-seq analyses.

Data File S5: Ingenuity pathway analysis for canonical pathways from single cell RNA-seq analyses.

Data File S6: Ingenuity pathway analysis for upstream regulating cytokines from single cell RNA-seq analyses.

Data File S7: Abundance of top respiratory microbes detected in each patient at the early and late timepoints.

Data File S8: Detailed clinical and demographic data for patients in this study.

## Data Availability

Host gene expression data are available under NCBI GEO accession number GSE168019 for bulk RNA-seq and GSE168018 for scRNA-seq. Raw microbial sequencing alignments are available from NCBI SRA under BioProject PRJNA704082. Code is available at https://github.com/bspeco/VAPinCOVID19.

## Author contributions

Conceptualization: CRL, AT, BSZ, AB, CD, SL, ER, AP, OR, CSC

Methodology: CRL, AT, BSZ, AB, CD, SL, ER, AJT, OR

Data acquisition: RG, AJ, PHS, TJD, BSZ, AB, NGJ, AD

Formal analysis: AT, BSZ, AB, CD, SL, ER, AJT, EM, JA

Investigation: BSZ, AB, CD, SS, CSC, DJE

Funding acquisition: CRL, CSC, JLD, DJE

Supervision: CRL, OR, NN, JLD, CSC, DJE

Writing - original draft: AT, BSZ, AB, CD, SL, ER, CRL

Writing - review & editing: All authors

## Acknowledgements

This study was performed with support from the National Institute of Allergy and Infectious Diseases-sponsored Immunophenotyping Assessment in a COVID-19 Cohort (IMPACC) Network. We gratefully appreciate support from Amy Kistler, PhD, Jack Kamm, PhD, Saharai Caldera, BS and Maira Phelps BS.

## Data and materials availability

Host gene expression data are available under NCBI GEO accession number GSE168019 for bulk RNA-seq and GSE168018 for scRNA-seq. Raw microbial sequencing alignments are available from NCBI SRA under BioProject PRJNA704082. Code used for RNA-seq and microbiome analyses is available at https://github.com/bspeco/VAPinCOVID19.

