## Supplemental Tables and Figures for "Impaired immune signaling and changes in the lung microbiome precede secondary bacterial pneumonia in COVID-19"

**Supplementary Table 1:** Clinical and demographic characteristics of patients with COVID-19 who did or did not develop VAP at the "early" and "late" timepoints, and control patients who were intubated without LRTI, included in the bulk RNA sequencing primary analysis.

| | **VAP (early timepoint)** | **No-VAP (early timepoint)** | **P-value (early - VAP vs No-VAP)** | **VAP (late timepoint)** | **No-VAP (late timepoint)** | **P-value (late - VAP vs No-VAP)** | **Controls (early timepoint)** | **P-value (+/- COVID-19)** |
|---|---|---|---|---|---|---|---|---|
| **N** | 4 | 8 | | 5 | 8 | | 8 | |
| **Age in years, median (IQR)** | 59.5 (56.0, 66.0) | 49 (40.5, 64.0) | 0.268 | 51.0 (48.0, 53.0) | 59.5 (47.0, 64.5) | 0.271 | 63.0 (36.0, 79.0) | 0.642 |
| **Female (%)** | 1 (25.0%) | 3 (37.5%) | 1.00 | 2 (40.0%) | 4 (50.0%) | 1.00 | 4 (50.0%) | 0.648 |
| **Race (%)** | | | | | | | | |
| Black | 0 (0.0%) | 0 (0.0%) | - | 0 (0.0%) | 0 (0.0%) | - | 0 (0.0%) | - |
| Asian | 0 (0.0%) | 0 (0.0%) | - | 0 (0.0%) | 1 (12.5%) | 1.00 | 1 (12.5%) | 0.400 |
| White | 0 (0.0%) | 2 (25.0%) | 0.515 | 1 (20.0%) | 2 (25.0%) | 1.00 | 7 (87.5%) | <0.01 |
| Other | 4 (100.0%) | 5 (62.5%) | 0.491 | 4 (80.0%) | 5 (62.5%) | 1.00 | 0 (0.0%) | <0.01 |
| Multiple | 0 (0.0%) | 1 (12.5%) | 1.00 | 0 (0.0%) | 0 (0.0%) | - | 0 (0.0%) | 1.00 |
| **Hispanic ethnicity (%)** | 3 (75.0%) | 4 (50.0%) | 0.576 | 4 (80.0%) | 4 (50.0%) | 0.565 | 1 (12.5%) | 0.070 |
| **Comorbidities (%)** | | | | | | | | |
| Autoimmune | 0 (0.0%) | 0 (0.0%) | - | 0 (0.0%) | 1 (12.5%) | 1.00 | 2 (25.0%) | 0.147 |
| Cancer | 0 (0.0%) | 0 (0.0%) | - | 1 (20.0%) | 0 (0.0%) | 0.385 | 1 (12.5%) | 0.400 |
| COPD/Asthma | 0 (0.0%) | 2 (25.0%) | 0.515 | 2 (40.0%) | 1 (12.5%) | 0.510 | 0 (0.0%) | 0.495 |
| DM | 4 (100.0%) | 3 (37.5%) | 0.081 | 3 (60.0%) | 4 (50.0%) | 1.00 | 3 (37.5%) | 0.650 |
| HTN | 3 (75.0%) | 1 (12.5%) | 0.067 | 2 (40.0%) | 1 (12.5%) | 0.510 | 4 (50.0%) | 0.648 |
| Obesity | 4 (100.0%) | 5 (62.5%) | 0.491 | 4 (80.0%) | 4 (50.0%) | 0.565 | 3 (37.5%) | 0.167 |
| Solid organ transplant | 0 (0.0%) | 0 (0.0%) | - | 0 (0.0%) | 0 (0.0%) | - | 0 (0.0%) | - |
| **Baseline IS meds (%)** | 0 (0.0%) | 0 (0.0%) | - | 1 (20.0%) | 1 (12.5%) | 1.00 | 1 (12.5%) | 0.400 |
| **Treated with immunosuppressants (%)** | 2 (50.0%) | 3 (42.9%)* | 1.00 | 5 (100.0%) | 4 (57.1%)* | 0.205 | 2 (25.0%) | 0.633 |
| **Steroid days before sample, median (IQR)** | 1.5 (0.0, 3.5) | 0.0 (0.0, 1.5) | 0.343 | 9.0 (7.5, 16.5) | 0.0 (0.0, 4.0) | **0.013** | 0.0 (0.0, 0.5) | 0.497 |
| **Antibiotic use before sample (%)** | 4 (100%) | 8 (100%) | - | 5 (100%) | 8 (100%) | - | 7 (87.5%) | 0.400 |
| **Length of stay, days, median (IQR)** | 57.5 (44.5, 65.6) | 19.0 (17.0, 34.5) | **0.017** | 34.0 (28.0, 38.0) | 24.0 (17.0, 34.5) | 0.213 | 10.5 (6.0, 19.5) | <0.01 |
| **Duration of mechanical ventilation, days, median (IQR)** | 22.5 (19.0, 27.0) | 14.0 (5.0, 23.5) | 0.174 | 18.0 (7.0, 25.0) | 6.5 (2.5, 16.0) | 0.271 | 11.0 (11.0, 14.0) | 0.151 |
| **Vasopressors[a] (%)** | 4 (100.0%) | 7 (87.5%) | 1.00 | 5 (100.0%) | 8 (100.0%) | - | 4 (50.0%) | 0.023 |
| **ECMO[b] (%)** | 0 (0.0%) | 0 (0.0%) | - | 0 (0.0%) | 0 (0.0%) | - | 0 (0.0%) | - |
| **WHO Severity Score[c], median (IQR)** | 7 (7, 7.5) | 7 (7, 7) | 0.151 | 7 (7, 8) | 7 (7, 7) | 0.062 | - | - |
| **Mortality (%)** | 1 (25.0%) | 0 (0.0%) | 0.333 | 2 (40.0%) | 0 (0.0%) | 0.128 | 1 (12.5%) | 1.00 |
| **Days before VAP (median [range])** | 16.5 (15, 38) | - | - | 2 (2, 3) | - | - | - | - |
| **Days post intubation (median [range])** | 2 (1, 5) | 2 (1, 4) | 0.930 | 17 (8, 23) | 9 (6, 19) | 0.160 | 1 (1, 3) | 0.062 |

*Immunosuppressant use unknown for one No-VAP patient due to enrollment in a double-blinded clinical trial; N of 7 used
[a] Defined as any requirement for vasopressors during admission
[b] Defined as ever requiring ECMO during admission
[c] Defined as maximum value for World Health Organization Ordinal Scale for Clinical Improvement during admission
P-values represent comparisons of patients with or without VAP at the early or late timepoint, or all COVID-19 patients at the early timepoint compared to intubated controls. Statistical significance was determined using Fisher's exact test (categorical variables) or by Wilcoxon test (continuous variables). Abbreviations: COPD: chronic obstructive pulmonary disease; DM: diabetes mellitus; ECMO: extracorporeal membrane oxygenation; HTN: hypertension; IQR: interquartile range; IS: immunosuppressive; VAP: ventilator-associated pneumonia; WHO: World Health Organization

**Supplementary Table 2:** Clinical and demographic characteristics of patients with COVID-19 who do or do not develop VAP at the “early” and “late” timepoints who were included in the single cell RNA sequencing analysis.

| | VAP (early timepoint) | No-VAP (early timepoint) | P-value (early - VAP vs No-VAP) | VAP (late timepoint) | No-VAP (late timepoint) | P-value (late - VAP vs No-VAP) |
|---|---|---|---|---|---|---|
| **N** | 4 | 6 | | 4 | 4 | |
| **Age in years, median (IQR)** | 64.0 (50.5, 76.5) | 56.5 (47.0, 68.0) | 0.392 | 61.5 (37.5, 74.0) | 60.0 (50.5, 74.0) | 0.271 |
| **Female (%)** | 2 (50.0%) | 2 (33.3%) | 1.00 | 0 (0.0%) | 2 (50.0%) | 0.429 |
| **Race (%)** | | | | | | |
| Black | 0 (0.0%) | 1 (16.7%) | 1.00 | 0 (0.0%) | 1 (25.0%) | 1.00 |
| Asian | 0 (0.0%) | 0 | - | 0 (0.0%) | 0 (0.0%) | - |
| White | 0 (0.0%) | 2 (33.3%) | 0.467 | 1 (25.0%) | 1 (25.0%) | 1.00 |
| Other | 4 (100.00%) | 3 (50.0%) | 0.200 | 3 (75.0%) | 2 (50.0%) | 1.00 |
| **Hispanic ethnicity (%)** | 4 (100.0%) | 2 (33.3%) | 0.076 | 4 (100.0%) | 2 (50.0%) | 0.429 |
| **Comorbidities (%)** | | | | | | |
| Autoimmune | 0 (0.0%) | 0 (0.0%) | - | 0 (0.0%) | 0 (0.0%) | - |
| Cancer | 2 (50.0%) | 0 (0.0%) | 0.133 | 1 (25.0%) | 0 (0.0%) | 1.00 |
| COPD/Asthma | 0 (0.0%) | 2 (33.3%) | 0.467 | 1 (25.0%) | 2 (50.0%) | 1.00 |
| DM | 1 (25.0%) | 3 (50.0%) | 0.571 | 2 (50.0%) | 2 (50.0%) | 1.00 |
| HTN | 1 (25.0%) | 1 (16.7%) | 1.00 | 2 (50.0%) | 3 (75.0%) | 1.00 |
| Obesity | 1 (25.0%) | 3 (50.0%) | 0.571 | 2 (50.0%) | 3 (75.0%) | 1.00 |
| Solid organ transplant | 0 (0.0%) | 0 (0.0%) | - | 0 (0.0%) | 1 (25.0%) | 1.00 |
| **Baseline IS meds (%)** | 1 (25.0%) | 0 (0.0%) | 0.400 | 0 (0.0%) | 1 (25.0%) | 1.00 |
| **Treated with immunosuppressants (%)** | 4 (100.0%) | 2 (33.3%) | 0.076 | 4 (100.0%) | 3 (75.0%) | 1.00 |
| **Steroid days before sample, median (IQR)** | 1 (1, 7) | 0 (0, 1) | 0.170 | 5.5 (2.5, 6.5) | 8.5 (3.5, 10) | 0.240 |
| **Antibiotic use before sample (%)** | 4 (100.0%) | 6 (100.0%) | - | 4 (100.0%) | 4 (100.0%) | - |
| **Length of stay, days, median (IQR)** | 33.0 (25.5, 38.0) | 29.0 (18.0, 37.0) | 0.521 | 38.0 (28.5, 113.5) | 27.0 (23.5, 30.0) | 0.245 |
| **Duration of mechanical ventilation, days, median (IQR)** | 20.0 (14.0, 23.5) | 10.5 (7.0, 19.0) | 0.286 | 23.5 (14.5, 29.5) | 10.5 (7.0, 25.0) | 0.564 |
| **Vasopressors[a] (%)** | 4 (100.0%) | 6 (100.0%) | - | 4 (100.0%) | 4 (100.0%) | - |
| **ECMO[b] (%)** | 0 (0.0%) | 0 (0.0%) | - | 0 (0.0%) | 0 (0.0%) | - |
| **WHO Severity Score[c], median (IQR)** | 8 (7.5, 8) | 7 (7, 7) | **0.016** | 7 (7, 7.5) | 7 (7, 7) | 0.317 |
| **Mortality (%)** | 3 (75.0%) | 0 (0.0%) | **0.033** | 1 (25.0%) | 0 (0.0%) | 1.00 |
| **Days before VAP, median (range)** | 9.5 (8, 16) | - | - | 3 (2, 4) | - | - |
| **Days after intubation, median (range)** | 5.5 (1, 10) | 4.0 (2, 13) | 0.748 | 8.5 (7, 18) | 10.5 (4, 15) | 0.884 |

*Immunosuppressant use unknown for one No-VAP patient due to enrollment in a double-blinded clinical trial; N of 7 used

[a] Defined as any requirement for vasopressors during admission

[b] Defined as ever requiring ECMO during admission

[c] Defined as maximum value for World Health Organization Ordinal Scale for Clinical Improvement during admission

P-values represent comparisons of patients with or without VAP at the early or late timepoint, or all COVID-19 patients at the early timepoint compared to intubated controls. Statistical significance was determined using Fisher’s exact test (categorical variables) or by Wilcoxon test (continuous variables). Abbreviations: COPD: chronic obstructive pulmonary disease; DM: diabetes mellitus; ECMO: extracorporeal membrane oxygenation; HTN: hypertension; IQR: interquartile range; IS: immunosuppressive; VAP: ventilator-associated pneumonia; WHO: World Health Organization

**Supplementary Table 3:** Details of immunosuppressant use for patients included in the primary bulk and single-cell RNA sequencing analyses at both “early” and “late” timepoints.

| Patient ID | Analysis | Group | IS before sample | Immunosuppressant details | Steroid days |
|---|---|---|---|---|---|
| 1059 | Bulk RNA-seq | VAP-early | N | - | 0 |
| 1154 | Bulk RNA-seq | VAP-early | Y | Dexamethasone 6 mg daily | 4 |
| 1196 | Bulk RNA-seq | VAP-early | N | - | 0 |
| 1254 | Bulk RNA-seq | VAP-early | Y | Dexamethasone 6 mg daily | 3 |
| 1001 | Bulk RNA-seq | No-VAP-early | N | - | 0 |
| 1002 | Bulk RNA-seq | No-VAP-early | N | - | 0 |
| 1072 | Bulk RNA-seq | No-VAP-early | N | - | 0 |
| 1115 | Bulk RNA-seq | No-VAP-early | N | - | 0 |
| 1136 | Bulk RNA-seq | No-VAP-early | Y | Betamethasone 12 mg x 1 | 1 |
| 1158 | Bulk RNA-seq | No-VAP-early | Unknown* | Baricitinib vs placebo* | 0 |
| 1185 | Bulk RNA-seq | No-VAP-early | Y | Dexamethasone 6 mg daily | 2 |
| 1223 | Bulk RNA-seq | No-VAP-early | Y | Dexamethasone 6 mg daily | 2 |
| 227 | Bulk RNA-seq | Control | N | - | 0 |
| 413 | Bulk RNA-seq | Control | N | - | 0 |
| 414 | Bulk RNA-seq | Control | N | - | 0 |
| 462 | Bulk RNA-seq | Control | N | - | 0 |
| 473 | Bulk RNA-seq | Control | N | - | 0 |
| 477 | Bulk RNA-seq | Control | Y | Dexamethasone 0.5 mg daily, fludrocortisone 0.1 mg daily | 1 |
| 555 | Bulk RNA-seq | Control | N | | 0 |
| 563 | Bulk RNA-seq | Control | Y | Dexamethasone 10 mg x1, then 4 mg q6 hours | 5 |
| 1047 | Bulk RNA-seq | VAP-late | Y | Methylprednisolone 40 mg IV q6, tapered over 6 days to home dexamethasone 4 mg daily | 23 |
| 1154 | Bulk RNA-seq | VAP-late | Y | Dexamethasone 6 mg daily | 7 |
| 1172 | Bulk RNA-seq | VAP-late | Y | Dexamethasone 6 mg daily | 8 |
| 1196 | Bulk RNA-seq | VAP-late | Y | Dexamethasone 6 mg daily | 10 |
| 1233 | Bulk RNA-seq | VAP-late | Y | Hydrocortisone 100 mg IV q8, tapered over 1-7 days [#]; tocilizumab x1 | 1-7[#] |
| 1001 | Bulk RNA-seq | No-VAP-late | Y | Prednisone 50 mg x 1 | 1 |
| 1002 | Bulk RNA-seq | No-VAP-late | N | - | 0 |
| 1038 | Bulk RNA-seq | No-VAP-late | Y | Tocilizumab x 2 | 0 |

| | | | | | |
|---|---|---|---|---|---|
| 1072 | Bulk RNA-seq | No-VAP-late | N | - | 0 |
| 1115 | Bulk RNA-seq | No-VAP-late | N | - | 0 |
| 1158 | Bulk RNA-seq | No-VAP-late | Unknown* | Baricitinib vs placebo* | 0 |
| 1185 | Bulk RNA-seq | No-VAP-late | Y | Dexamethasone 6 mg daily | 8 |
| 1244 | Bulk RNA-seq | No-VAP-late | Y | Dexamethasone 6 mg daily | 7 |
| 1047 | scRNA-seq | VAP-early | Y | Methylprednisolone 40 mg IV q 6 hours, tapered over 6 days to home dexamethasone 4 mg daily | 9 |
| 1154 | scRNA-seq | VAP-early | Y | Dexamethasone 6 mg daily | 9 |
| 1161 | scRNA-seq | VAP-early | Y | Dexamethasone 10 mg x 1, then 6 mg daily; baricitinib vs placebo | 4 |
| 1357 | scRNA-seq | VAP-early | Y | Dexamethasone 6 mg daily | 1 |
| 1001 | scRNA-seq | No-VAP-early | N | - | 0 |
| 1002 | scRNA-seq | No-VAP-early | N | - | 0 |
| 1072 | scRNA-seq | No-VAP-early | N | - | 0 |
| 1115 | scRNA-seq | No-VAP-early | N | - | 0 |
| 1158 | scRNA-seq | No-VAP-early | Unknown* | Baricitinib vs placebo* | 0 |
| 1271 | scRNA-seq | No-VAP-early | Y | Dexamethasone 6 mg daily; canakinumab vs placebo | 10 |
| 1154 | scRNA-seq | VAP-late | Y | Dexamethasone 6 mg daily | 7 |
| 1233 | scRNA-seq | VAP-late | Y | Hydrocortisone 100 mg IV q8 hours, tapered over 1-7 days [#]; tocilizumab x1 | 1-7[#] |
| 1264 | scRNA-seq | VAP-late | Y | Dexamethasone 6 mg daily | 10 |
| 1357 | scRNA-seq | VAP-late | Y | Dexamethasone 6 mg daily | 6 |
| 1072 | scRNA-seq | No-VAP-late | N | - | 0 |
| 1271 | scRNA-seq | No-VAP-late | Y | Dexamethasone 6 mg daily; canakinumab vs placebo | 10 |
| 1290 | scRNA-seq | No-VAP-late | Y | Dexamethasone 6 mg daily; canakinumab vs placebo | 7 |
| 1320 | scRNA-seq | No-VAP-late | Y | Dexamethasone 6 mg daily x 10 days, then home prednisone 5mg daily; home tacrolimus; home mycophenolate mofetil | 12 |

*Unknown due to enrollment in double-blinded clinical trial of baricitinib vs placebo. [#] Exact duration unknown to due to incomplete records on transfer from an outside facility. Abbreviations: IS = immunosuppressive.

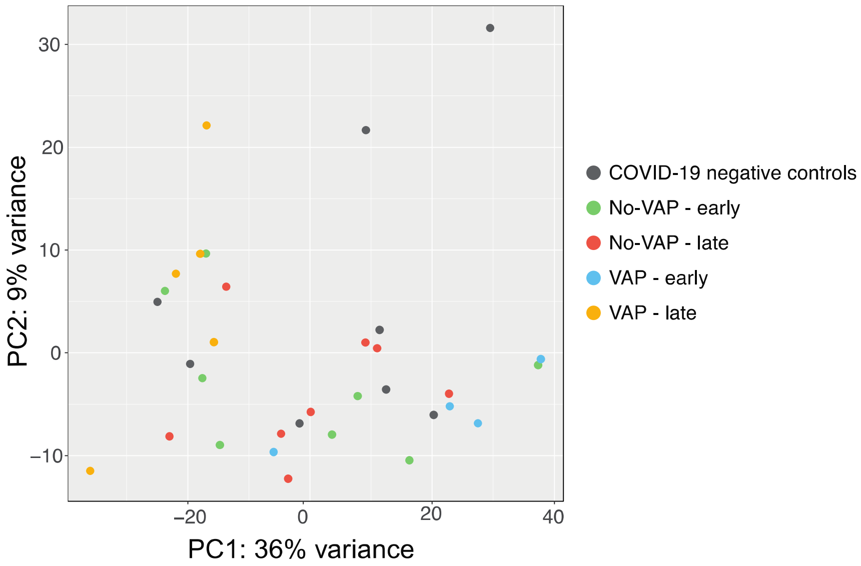


**Supplementary Figure 1:** Principal component analysis (PCA) plot of all samples included in bulk RNA-seq analyses.

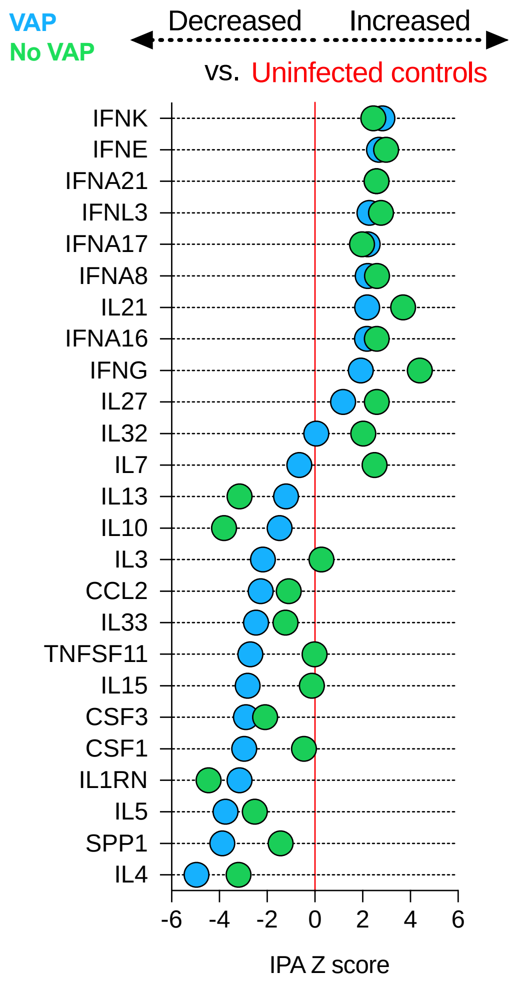


**Supplementary Figure 2:** Regulation of cytokines at the "early" timepoint with respect to a baseline of uninfected, intubated controls. IPA upstream regulator analysis was based on differential gene expression analyses of patients with VAP vs controls and No-VAP patients vs controls. Cytokines were selected from the IPA results if they had a Z-score absolute value > 2 and an overlap P-value < 0.05 in both of the comparisons (VAP vs controls and No-VAP vs controls). All cytokines are tabulated in Supplementary Data File 3.

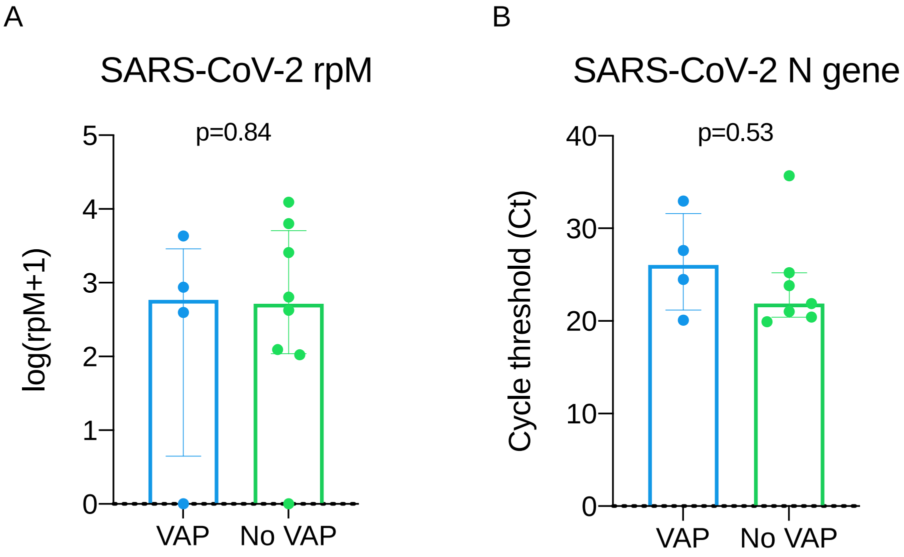


**Supplementary Figure 3:** SARS-CoV-2 viral load in VAP and No-VAP patients at the “early” timepoint. Viral load was quantified by **A)** reads per million (rpM) from bulk RNA-seq and **B)** cycle threshold (Ct) values by qPCR for the SARS-CoV-2 N gene. The Ct value for one sample in the No-VAP group was not determined. Bar plots represent the median with interquartile range. Statistical significance was determined by Mann-Whitney tests.

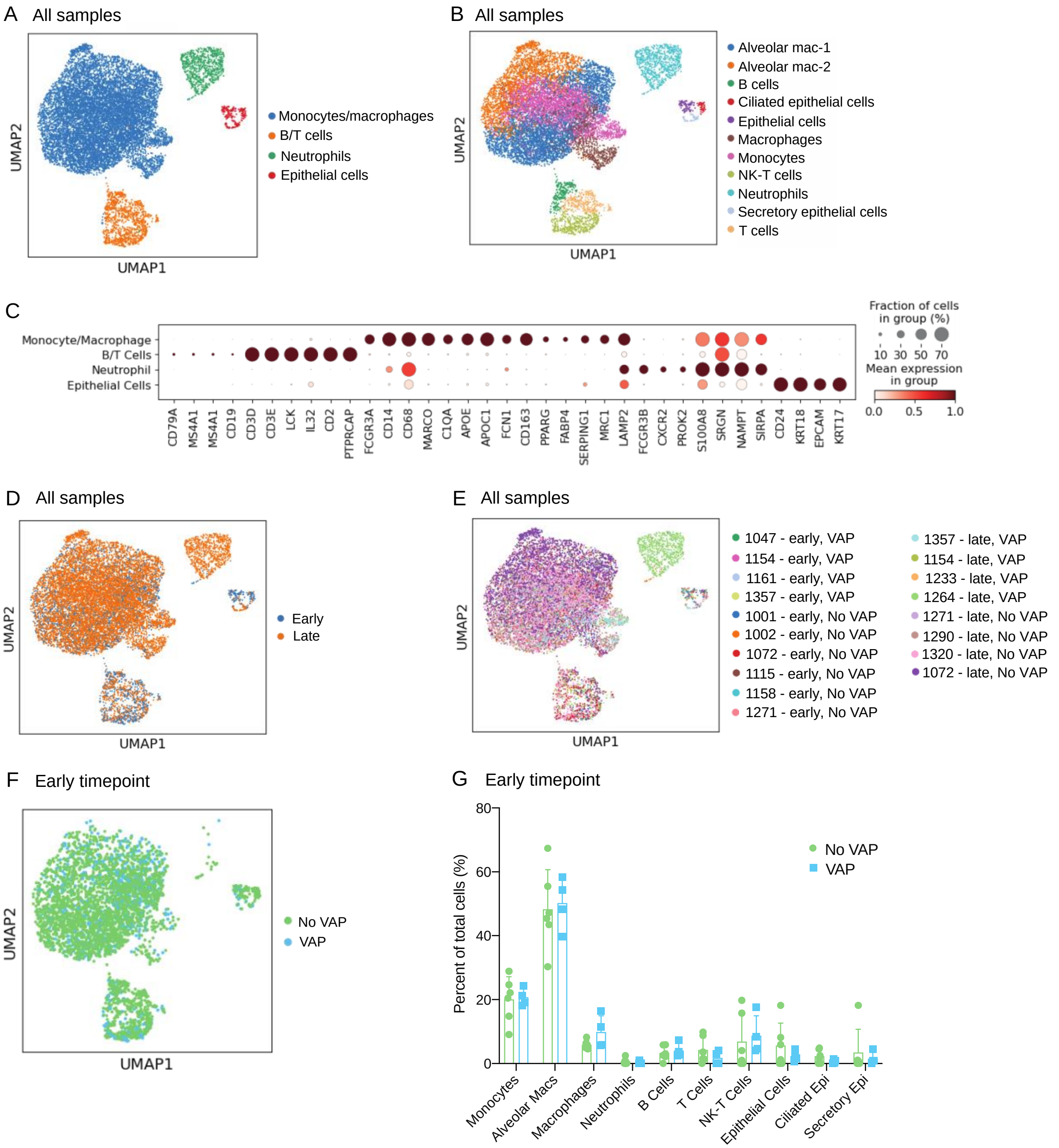


**Supplementary Figure 4:** UMAPs of all single cell RNA-seq samples, annotated by **(A)** low-resolution cell type and **(B)** high-resolution cell type. **(C)** Expression of marker genes used for annotation. UMAPs of all samples annotated by **(D)** timepoint and **(E)** sample. **(F)** UMAP of all samples at the “early” timepoint, annotated by VAP status. **(G)** Cell type proportions in VAP and No-VAP samples at the “early” timepoint. Mann-Whitney tests were conducted to determine statistical significance. None of the cell types reached statistical significance with $P < 0.05$.

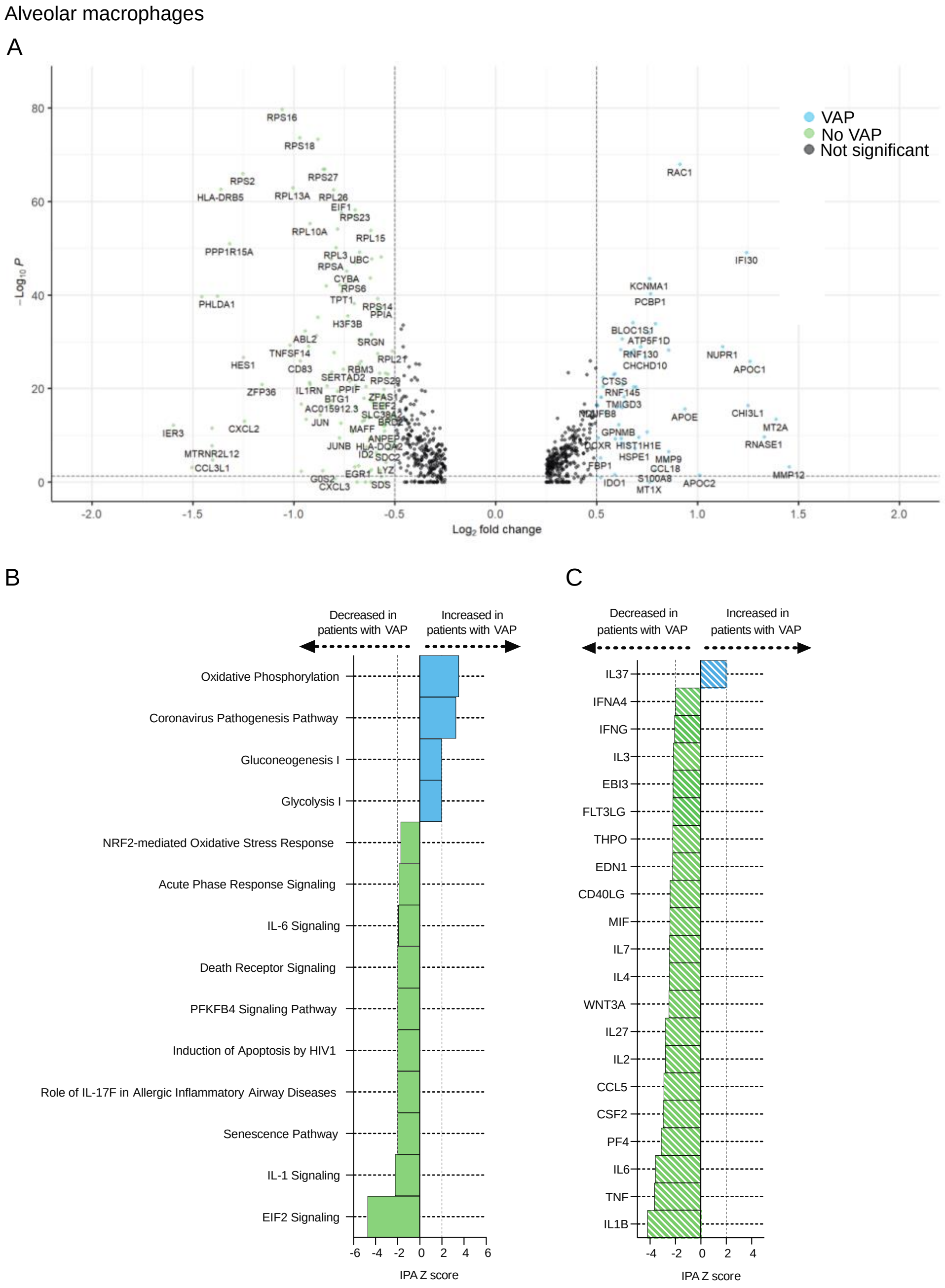


**Supplementary Figure 5: (A)** Volcano plot displaying the differentially expressed genes between VAP and No-VAP patients in alveolar macrophages at the "early" timepoint. **(B-C)** Ingenuity Pathway Analysis (IPA) of (B) canonical pathways and (C) upstream cytokine activation states based on differential gene expression in alveolar macrophages of patients who developed VAP versus those who did not, including genes with adjusted P-values < 0.05. Only significant pathways and cytokines (IPA Z-score absolute value > 2 and overlap P-value < 0.05) are depicted. All pathways are provided in Supplementary Data File 5.

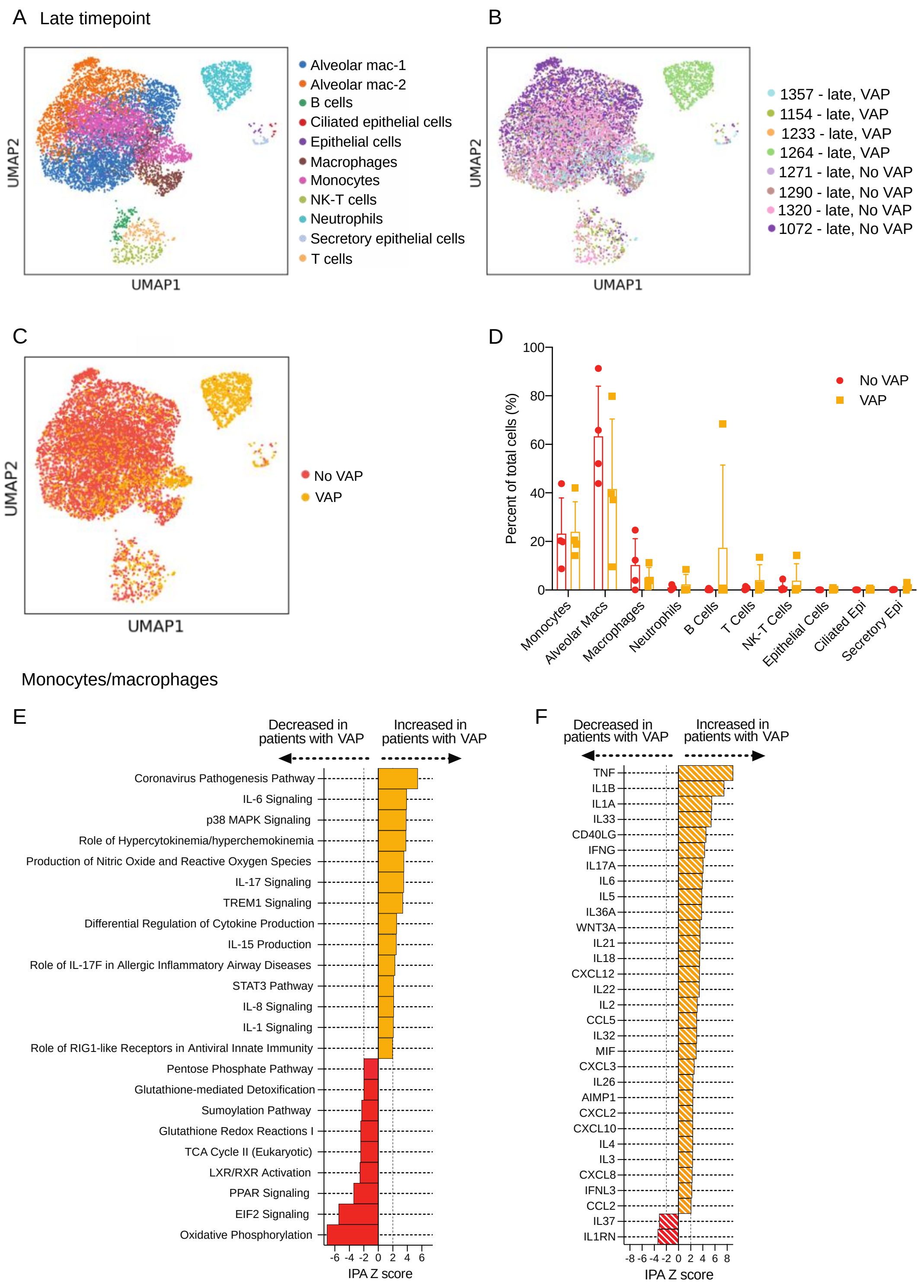


**Supplementary Figure 6:** UMAP of single cell RNA-seq samples at the "late" timepoint, annotated by **(A)** cell type, **(B)** sample, or **(C)** VAP status. **(D)** Cell type proportions in VAP and No-VAP samples at the "late" timepoint. Mann-Whitney tests were conducted to determine statistical significance. None of the cell types reached statistical significance with $P < 0.05$. **(E-F)**

Ingenuity Pathway Analysis (IPA) of (E) canonical pathways and (F) upstream cytokine activation states based on differential gene expression in monocytes/macrophages of patients who developed VAP versus those who did not at the "late" timepoint, including genes with adjusted P-values < 0.05. Only significant pathways (IPA Z-score absolute value > 2 and overlap P-value <0.05) are depicted. All pathways are provided in Supplementary Data File 5.

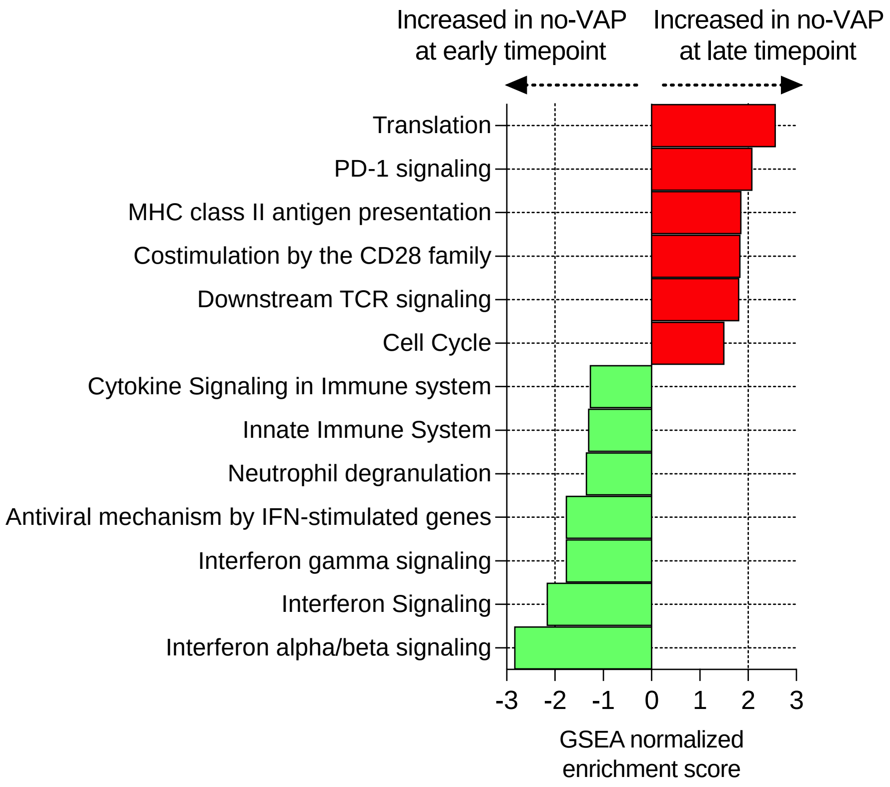


**Supplementary Figure 7**: Gene set enrichment analysis (GSEA) comparing patients who did not develop VAP at the “early” (green) versus “late” (red) timepoints. GSEA results were considered significant with an adjusted P-value < 0.05. All pathways are provided in Supplementary Data File 2.

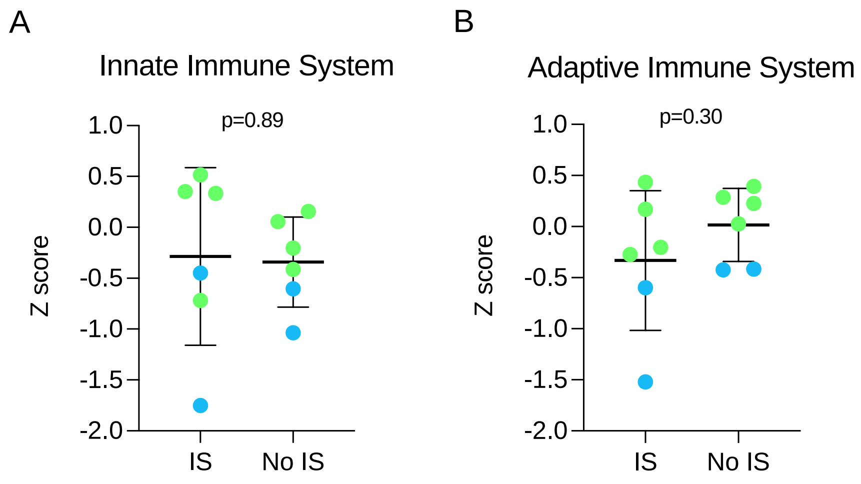


**Supplementary Figure 8:** Pathway expression for samples at the “early” timepoint, comparing patients who were treated with immunosuppressants before sample collection and those who were not. Pathway Z-scores for **(A)** the innate immune system pathway and **(B)** the adaptive immune system pathway were calculated by averaging Z-scores for the top 20 leading edge genes of each pathway. Samples from VAP patients are shown in blue, No-VAP patients are shown in green. IS = treated with immunosuppressants. Plots represent the mean with standard deviation. Statistical significance was determined by t-test.

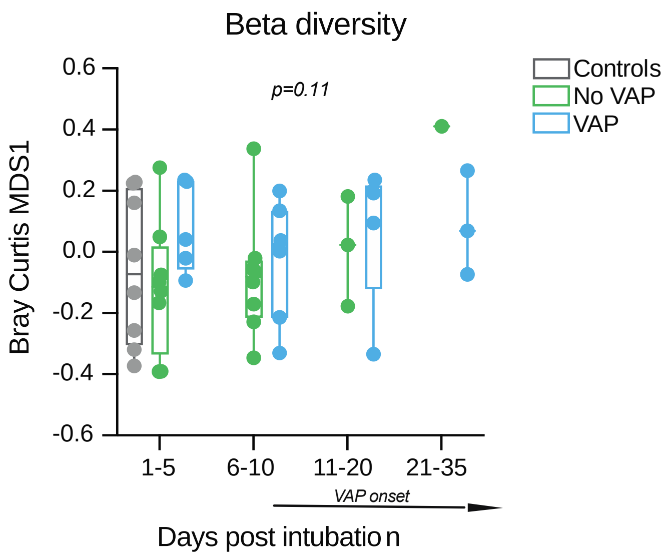


**Supplementary Figure 9:** Lung microbiome β–diversity (Bray Curtis Index, NMDS scaling) in COVID-19 patients with relation to VAP development over time by days since intubation**.**

**Supplementary Data Files**

**Data File S1:** Differentially expressed genes from bulk RNA-seq analyses.

**Data File S2:** Gene set enrichment analysis results from bulk RNA-seq analyses. NES = Normalized enrichment score.

**Data File S3:** Ingenuity pathway analysis for upstream regulating cytokines from bulk RNA-seq analyses.

**Data File S4:** Differentially expressed genes from single cell RNA-seq analyses.

**Data File S5:** Ingenuity pathway analysis for canonical pathways from single cell RNA-seq analyses.

**Data File S6:** Ingenuity pathway analysis for upstream regulating cytokines from single cell RNA-seq analyses.

**Data File S7:** Abundance of top respiratory microbes detected in each patient at the early and late timepoints.

**Data File S8:** Detailed clinical and demographic data for patients in this study.

**Supplementary Appendix:** COMET Consortium Member list.
